# Mental health conditions and COVID-19 vaccine outcomes: a scoping review

**DOI:** 10.1101/2023.10.27.23297663

**Authors:** Ru Jia, Carol Coupland, Yana Vinogradova, Nadeem Qureshi, Emma Turner, Kavita Vedhara

## Abstract

**Background:** The COVID-19 pandemic has had a profound impact on the mental health of people worldwide. Mental health also impacts on physical health. In the context of viral illnesses, viral challenge studies have shown that indices of mental health are associated with susceptibility to viral infections, including coronaviruses. Research conducted during the pandemic has shown that people with a history of mental health conditions were at increased risk of infection, hospitalisation, and mortality. However, the relationship between mental health conditions and vaccine outcomes such as vaccine intentions, uptake, and vaccine breakthrough is not yet well-understood.

**Methods:** We conducted a systematic search on the topics of COVID-19 vaccine intentions, vaccine uptake, and vaccine breakthrough, in relation to mental health conditions, in four databases: PubMed, MEDLINE, SCOPUS, and PsychINFO, as well as the publication lists of Clinical Practice Research Datalink (CPRD), The Health Improvement Network (THIN), OpenSAFELY, and QResearch. Inclusion criteria focus on studies reporting either of the aforementioned COVID-19 vaccine outcomes among people with mental health conditions.

**Results:** Thirty-three out of 251 publications met our inclusion criteria for this review. Overall, the evidence is inconclusive regarding the level of intention to accept the COVID-19 vaccine among people with mental health conditions. However, people with mental health conditions were more likely to have lower uptake of the COVID-19 vaccine, compared to people without. Common barriers to COVID-19 vaccine uptake include concerns about the safety, effectiveness, and side effects of the vaccines. Limited evidence also suggests that vaccine breakthrough may be a particular risk for those with substance use disorder.

**Conclusions:** Our findings revealed a possible intention-behaviour gap for receiving the COVID-19 vaccine among people with mental health conditions, yielding interventions to encourage vaccine uptake in this population. There is also the need to enhance our understanding of COVID-19 vaccine breakthrough in people with mental health conditions.

## Introduction

The COVID-19 pandemic resulted in more than 700 million cases and 6 million deaths globally (WHO, n.d.). Vaccines against SARS-CoV-2 were developed to reduce the spread of the virus and morbidity and mortality caused by COVID-19. The UK was the first country to approve a COVID-19 vaccine and started a vaccine roll-out program in December 2020 (Authority of the House of Commons, 2022). By autumn 2022, approximately 151 million vaccinations were given to the UK population (Majeed et al., 2022). However, while the mass vaccination roll-out was necessary in the pandemic, certain factors determine whether vaccines can realise their full potential. Arguably, the two most salient of these are vaccine uptake i.e., an individual’s willingness to be vaccinated and vaccine effectiveness i.e., the ability for vaccines to protect against infection and adverse outcomes.

One factor that may influence both vaccine uptake and vaccine effectiveness is mental health. Pre-pandemic the evidence suggested that people with mental illness might be less likely to engage in preventive healthcare such as blood pressure monitoring and vaccinations, compared with people without mental illness (Lord et al., 2010). Evidence during the pandemic seems to show a mixed picture. For example, a study in early 2021 found higher COVID-19 vaccine uptake (i.e., receiving at least one dose) among people with mood disorders (e.g., major depression and bipolar disorder) in certain areas in the UK, but lower COVID-19 vaccine uptake among individuals with psychotic disorders (e.g., schizophrenia), compared with people without mental illness (Hassan et al., 2022). Another study, conducted in the same year, found that the vaccination rate among people with mental health conditions (11%) was lower than the vaccination rate among other residents (40%) in Wuhan area (Huang et al., 2021). Reviews of the literature on vaccine hesitancy appear to confirm that the relationship between vaccine uptake and mental health varies according to the nature of the mental health problem (Farcas et al., 2022). However, attempts to review and synthesise this evidence have been limited to studies capturing the intention (or not) to be vaccinated, rather than actual uptake.

There is also potential for mental health to influence the effectiveness of COVID-19 vaccines. Mental health can dysregulate the immune system and thus can impair the immune response to vaccination (Cohen, 2021; O’Connor et al., 2021; Vedhara et al., 1999). For example, viral and vaccine challenge studies have shown that depression and stress are associated with poorer antibody responses to vaccinations, as well as increased risk of infection and symptomatic illness (Cohen, 2021; Cohen et al., 1998; Phillips et al., 2006; Vedhara et al., 1999).

Evidence from the pandemic suggests that this relationship between mental health and impaired responses to respiratory infections may also occur in the context of COVID-19. For example, observational data have shown that greater psychological distress was significantly associated with subsequent self-reported SARS-CoV-2 infection, together with an increased number of, and more severe, symptoms (Ayling et al., 2022). Similarly, Yang and colleagues reported that people with previous mental health conditions were more likely to get SARS-CoV-2 infection, be hospitalised, and to die from COVID-19 (Yang et al., 2020). However, no study, to our knowledge, has reviewed current evidence on the association between mental health conditions and COVID-19 vaccine breakthrough (i.e., evidence of SARS-CoV-2 infection after vaccination).

Thus, in summary, there is evidence to suggest that mental health conditions are associated with greater vaccine hesitancy, impaired immune responses to vaccination and greater risk of respiratory infections. In the context of COVID-19, the emerging evidence suggests that the relationship with vaccine hesitancy may vary between different mental health conditions; the relationship with vaccine uptake is less clear and while poorer COVID-19 outcomes have been associated with mental health conditions, the evidence on the relationship with vaccine effectiveness is limited. In an effort to bring this literature together, we present here results from a scoping review in which we have examined the evidence exploring the relationship between indices of mental health (past and current) and vaccine hesitancy (i.e., behavioural intentions), vaccine uptake (behavioural response) and vaccine effectiveness as measured by vaccine breakthrough i.e., evidence of infection following vaccination. An understanding of this literature will help to inform whether and in what ways people with mental health conditions need to be supported to reduce their risk of COVID-19 infection.

## Methods

### Population

The population of interest was people with mental health conditions. Consistent with previous work, mental health conditions included depression, anxiety, stress-related disorder, substance misuse, psychotic disorders, schizophrenia, and bipolar disorder (Thompson et al., 2022; Yang et al., 2020). We included studies involving people with a history of mental health difficulties as well as those arising during the pandemic. No restrictions were made for other characteristics, including age, gender, health conditions etc.

### Outcomes

The outcomes of interest were intention to accept or decline a COVID-19 vaccination when offered, COVID-19 vaccine uptake, and COVID-19 vaccine breakthrough. A COVID-19 vaccine breakthrough infection is defined according to CDC: “the detection of SARS-CoV-2 RNA or antigen in a respiratory specimen collected from a person ≥14 days after receipt of all recommended doses of an FDA-authorized COVID-19 vaccine” (Birhane et al., 2021).

### Search strategy

Searches for relevant studies were conducted in PubMed, MEDLINE, SCOPUS, and PsychINFO. To include potentially eligible population-based studies, the CPRD, The Health Improvement Network (THIN), OpenSAFELY, and QResearch lists of publications were also searched. To include articles that may be under review, the electronic pre-print service MedRxiv was also searched. The search paradigm was based on the following combination: (*mental, major* [MeSH terms]) AND (*COVID-19 vaccine* [MeSH terms]). To ensure our searches were comprehensive, the term *mental* was replaced with psychiatric, schizophrenia, psychotic, bipolar disorder, mood disorder, major depressive disorder, anxiety disorder, personality disorder, eating disorder, alcohol abuse, alcohol misuse, substance abuse, and substance misuse. We limited our searches to publications in English language. We did not limit the searches by time period.

### Eligibility criteria

Only observational studies were included. Eligible studies had to report at least one of the following: (1) intentions towards COVID-19 vaccine (i.e., intended willingness to accept or decline a vaccination when offered) among people with mental health conditions, and in comparison with people without mental health conditions where applicable; (2) COVID-19 vaccine uptake rate among people with mental health conditions, and in comparison with people without mental health conditions where applicable; (3) COVID-19 vaccine breakthrough (i.e., evidence of SARS-CoV-2 infection after vaccination) among people with mental health conditions. For mixed-method studies, only the observational part of the study was included. Only articles reporting results from original observational studies were included. Qualitative studies, reviews, protocols, commentary or opinion pieces, editorial letters and posters were excluded.

### Barriers to COVID-19 vaccine uptake

We were also interested in the potential predictors to COVID-19 vaccine intention, and vaccine uptake. However, predictors of COVID-19 vaccine intention have been systematically reviewed in previous work (see Farcas et al., 2022; Terry et al., 2022; Q. Wang et al., 2021). Demographic predictors (e.g., age, gender, ethnicity, education, socioeconomic status, etc) of vaccine uptake in general have also been reviewed previously (see Galanis et al., 2021). Therefore, in the current review, where studies reported on psychological barriers to COVID-19 vaccine uptake (e.g., beliefs, trust, etc.), we summarised these findings.

## Results

### Selection of studies

The database search was conducted on 9^th^ November 2022 and returned 251 initial results. Of these, 197 studies were excluded after title and abstract screening and 21 studies were excluded after full-text screening (Figure 1). The main reasons for exclusion include the article being a comment piece or review of literature or intervention etc. (k=51, 23%); studies not conducted among people with mental health conditions (k=42, 28%), and not investigating the outcomes of interest (k=16, 8%). Therefore, 33 studies were included in this scoping review.

**Figure 1.**
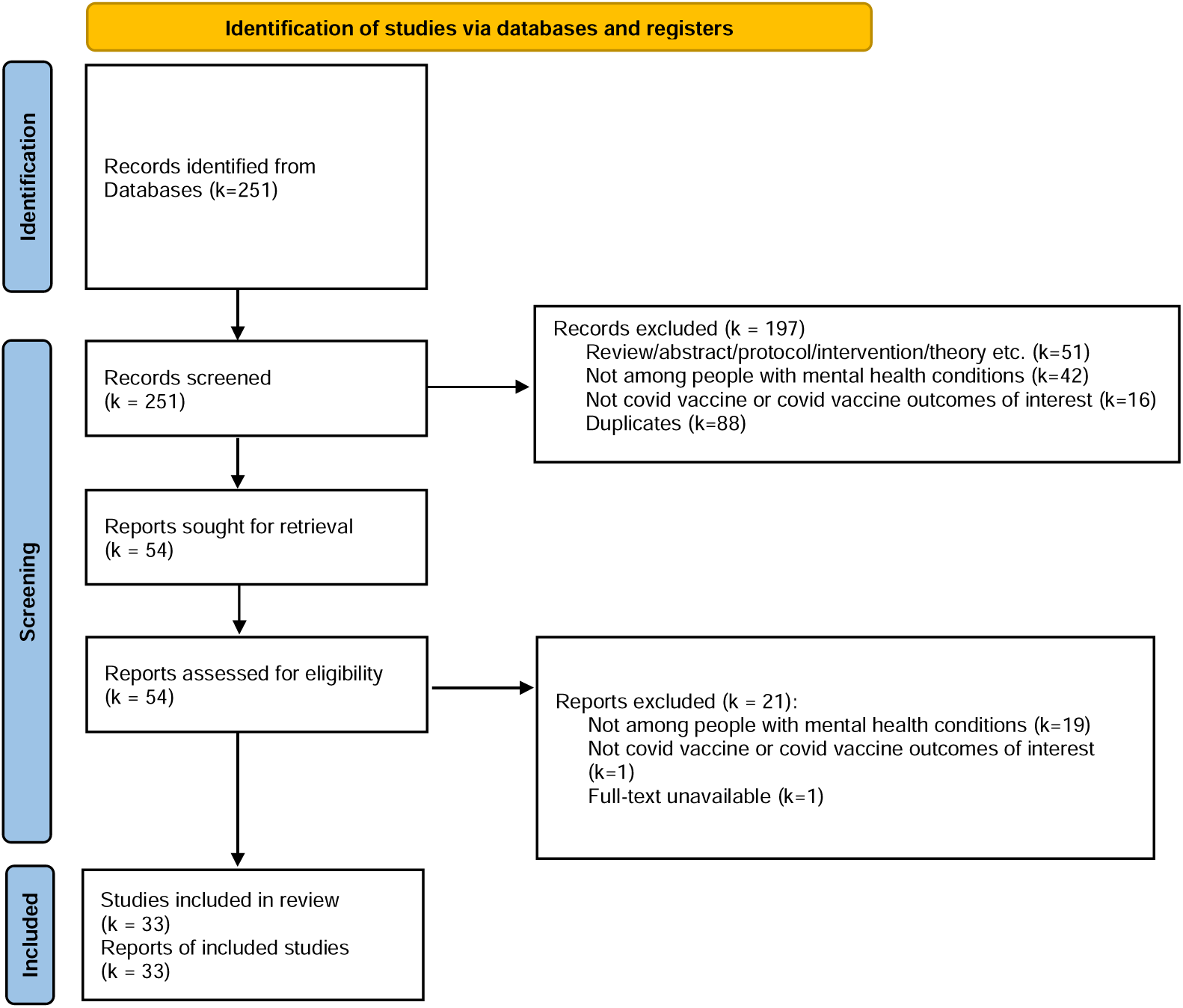
Flow diagram for the scoping review

### Overview of study characteristics

The 33 studies included in this review were conducted across 13 countries and regions including North America (e.g., Canada and the United States), Europe (e.g., United Kingdom, Belgium, France, Denmark), Asia (e.g., Mainland China, Japan, India, Taiwan), and the Middle East (e.g., Israel and Qatar). Of the 33 studies, 22 used survey designs including 6 longitudinal survey studies. Ten studies collected data from electronic health or registration records, including one study linking survey and electronic health record data. Two studies examined vaccine outcomes among patients within psychiatric hospitals through healthcare screening and assessment. Populations investigated by these studies included the general public or population-representative cohorts, and more specific cohorts for instance veterans, psychiatric patients, adolescents with attention-deficit/hyperactivity disorder (ADHD), and youth who are 2-spirit, lesbian, gay, bisexual, transgender, queer, and questioning (2SLGBTQ+) and experiencing homelessness, etc. One study investigated the intentions to vaccinate children with ADHD among their caregivers (Tsai et al., 2022). The sizes of the studies range from 62 to 57.9 million.

### Measurement of mental health conditions

Twenty-one of the 33 reviewed studies identified patients with mental health conditions through clinical diagnoses obtained from medical records (Bai et al., 2021; Cai et al., 2022; Curtis, Inglesby, MacKenna, et al., 2022; Curtis, Inglesby, Morton, et al., 2022; Eyllon et al., 2022; Gibbon et al., 2021; Hassan et al., 2022; Jefsen et al., 2021; Mazereel et al., 2021; Moeller et al., 2021; Nilsson et al., 2022; Nishimi, Neylan, et al., 2022; Qin et al., 2022; Raffard et al., 2022; Ren et al., 2021; Shkalim Zemer et al., 2022; Sullivan et al., 2022; Tsai et al., 2022; Tzur Bitan et al., 2022; Uvais, 2022; L. Wang et al., 2022).

The recorded mental health conditions included depressive disorder, bipolar disorder, schizophrenia, ADHD, PTSD, generalized anxiety disorder, and substance use disorder, etc. One study used records of prescription of psychotropic drugs (i.e., anxiolytics, antipsychotics, hypnotics, and antidepressants) as an indication of patients’ mental health conditions (Murphy et al., 2022). Six studies asked participants to self-report whether they have been diagnosed with mental health conditions (Afifi et al., 2021; Balut et al., 2021; Dvorsky et al., 2022; Huang et al., 2021; Paul & Fancourt, 2022; Roberts et al., 2022). Five other studies measured self-reported anxiety, depression, substance use, and posttraumatic stress disorder (PTSD) through surveys such as the Physician Health Questionnaire (PHQ-9, Kroenke et al., 2010), the 7-item Generalized Anxiety Disorder Questionnaire (GAD-7, Spitzer et al., 2006), and the 20-item posttraumatic stress disorder Checklist-5 (PCL-5, Blevins et al., 2015; Abramovich et al., 2022; Khaled et al., 2021; Nguyen et al., 2022; Nishimi et al., 2022; Sekizawa et al., 2022).

### Mental health conditions and COVID-19 vaccine intentions

Eighteen of the 33 reviewed studies reported on the association between the presence of mental health conditions and willingness or hesitancy to accept the COVID-19 vaccine or booster vaccine (Abramovich et al., 2022; Afifi et al., 2021; Bai et al., 2021; Cai et al., 2022; Dvorsky et al., 2022; Eyllon et al., 2022; Huang et al., 2021; Jefsen et al., 2021; Khaled et al., 2021; Nguyen et al., 2022; Nishimi, Borsari, et al., 2022; Paul & Fancourt, 2022; Qin et al., 2022; Ren et al., 2021; Roberts et al., 2022; Sekizawa et al., 2022; Sullivan et al., 2022; Tsai et al., 2022). The sizes of the studies range from 92 to 77104 (see Table 1). Participants in these studies were asked to indicate their intentions once the COVID-19 vaccines became available. Seventeen of the 18 studies reported on people with current mental health conditions. In the remaining study, no distinction was made between current and history of mental health conditions (Roberts et al., 2022). Therefore, we do not distinguish between current and history of mental health conditions in the following reporting.

**Table 1.**
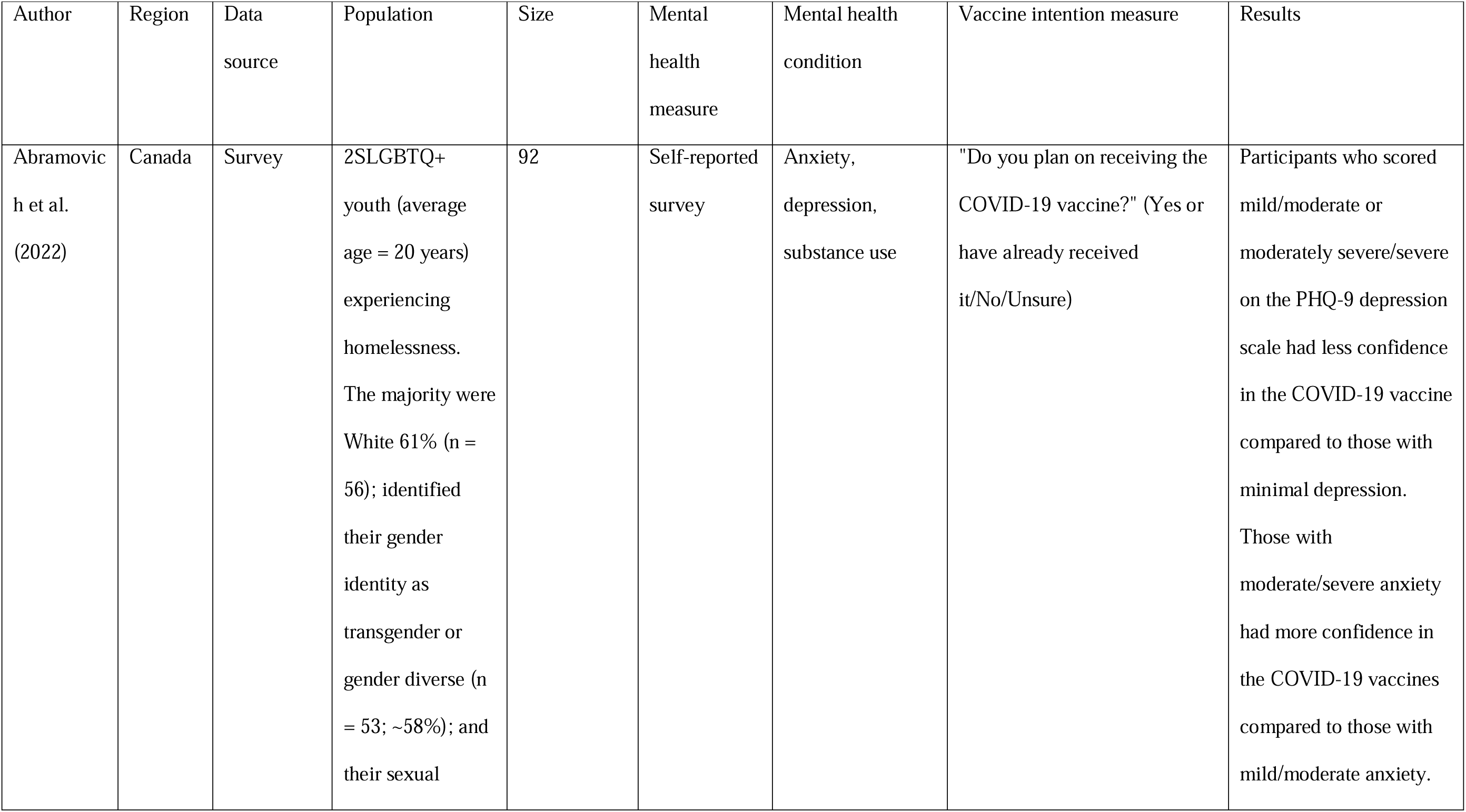
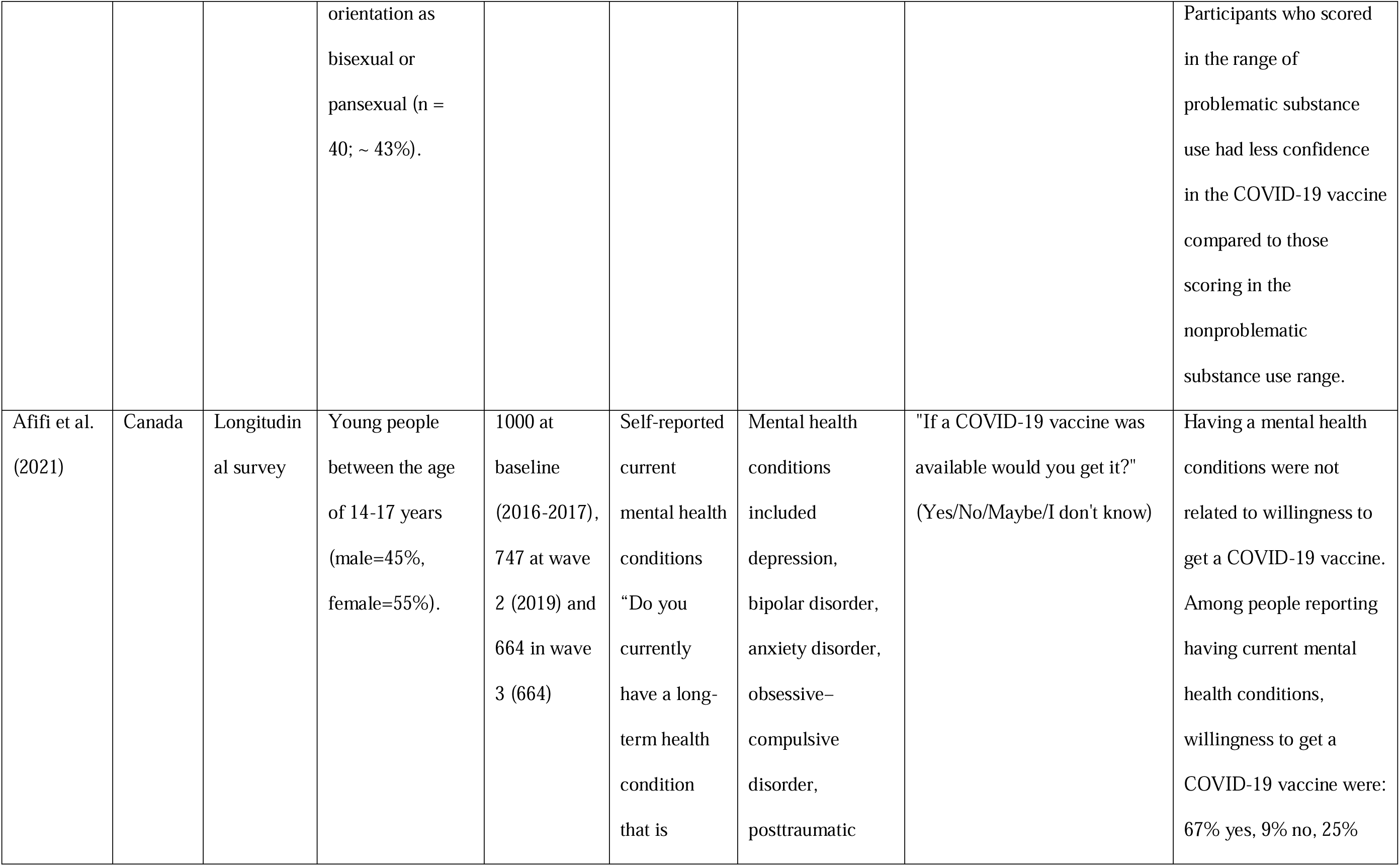
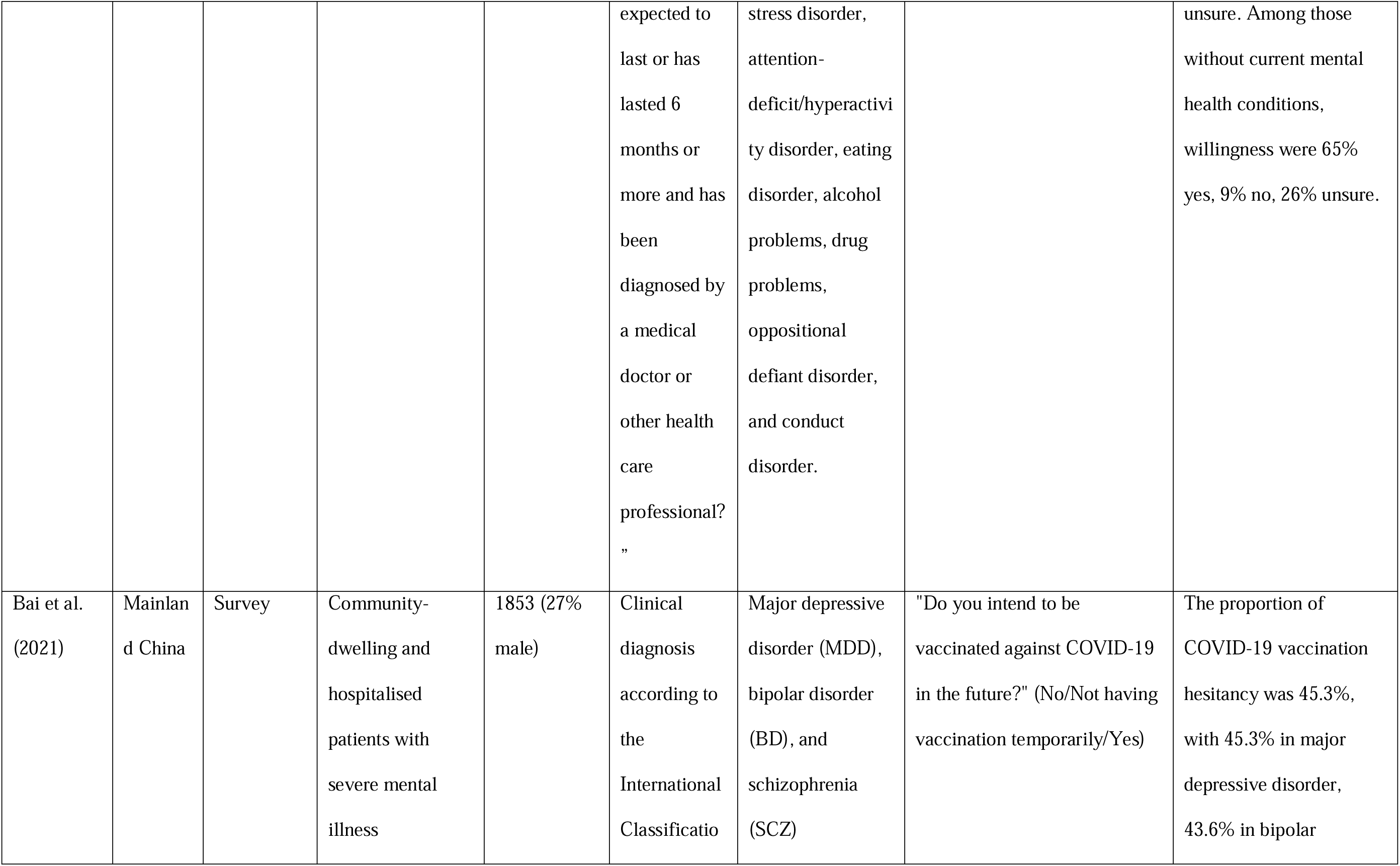
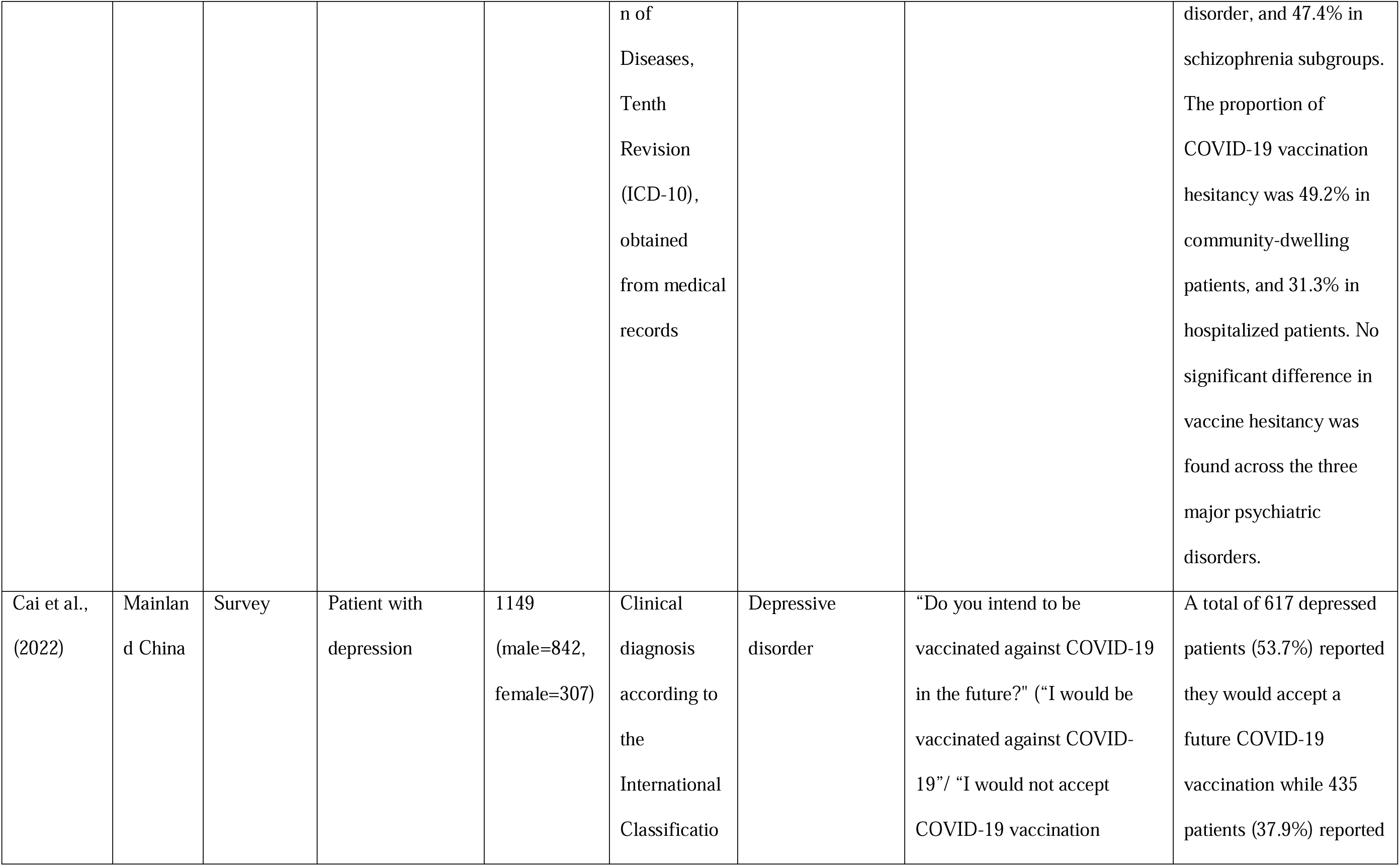
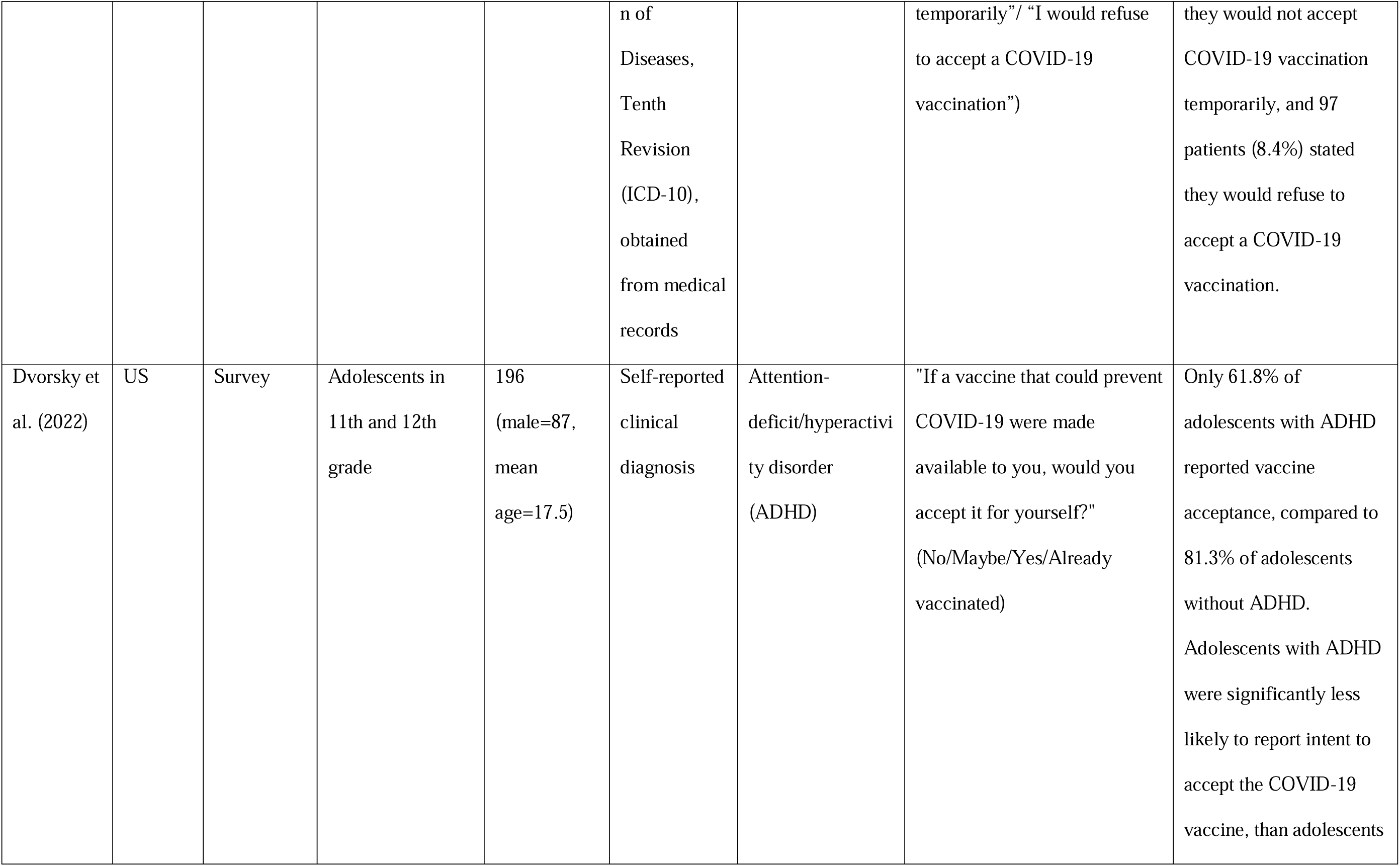
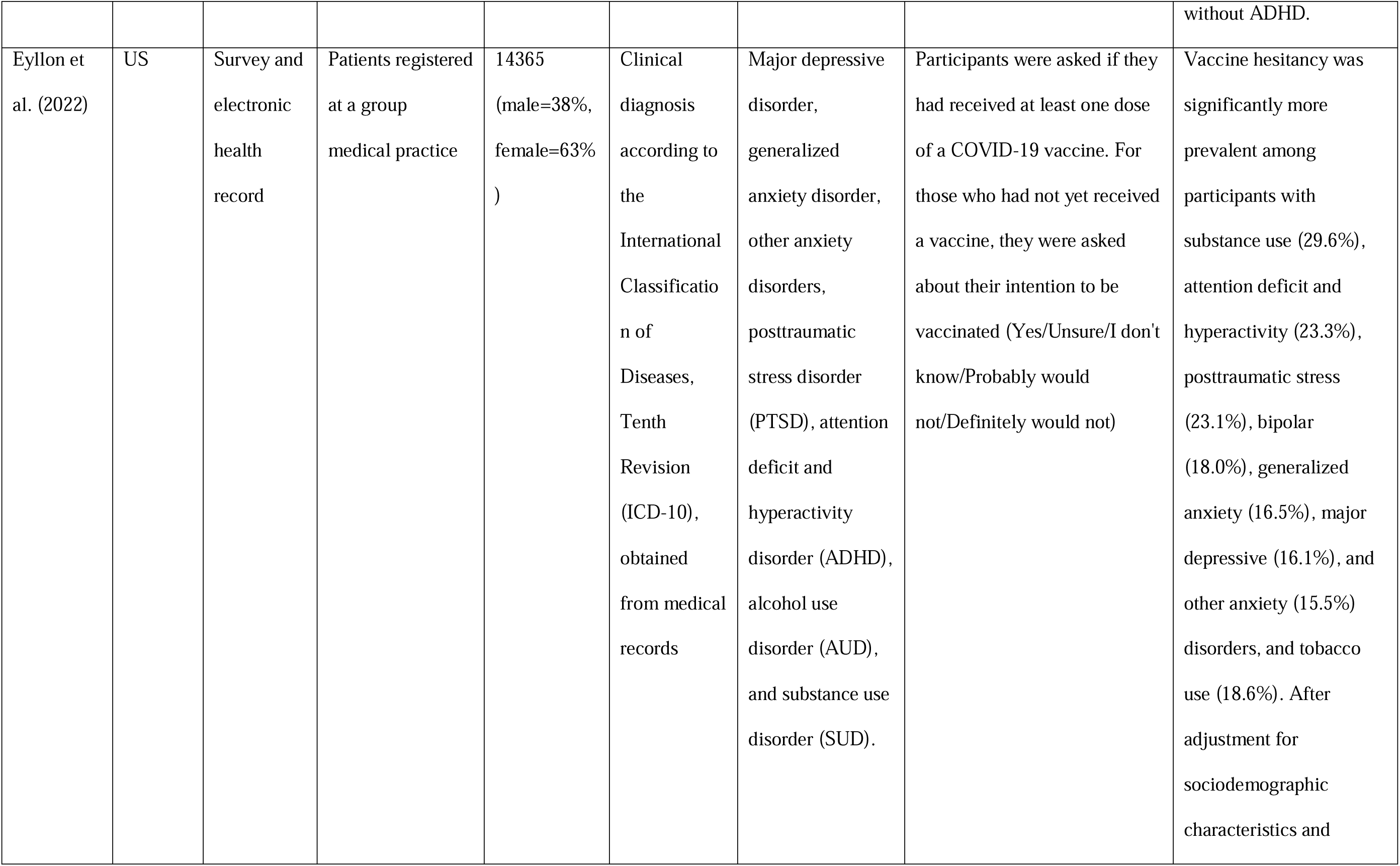
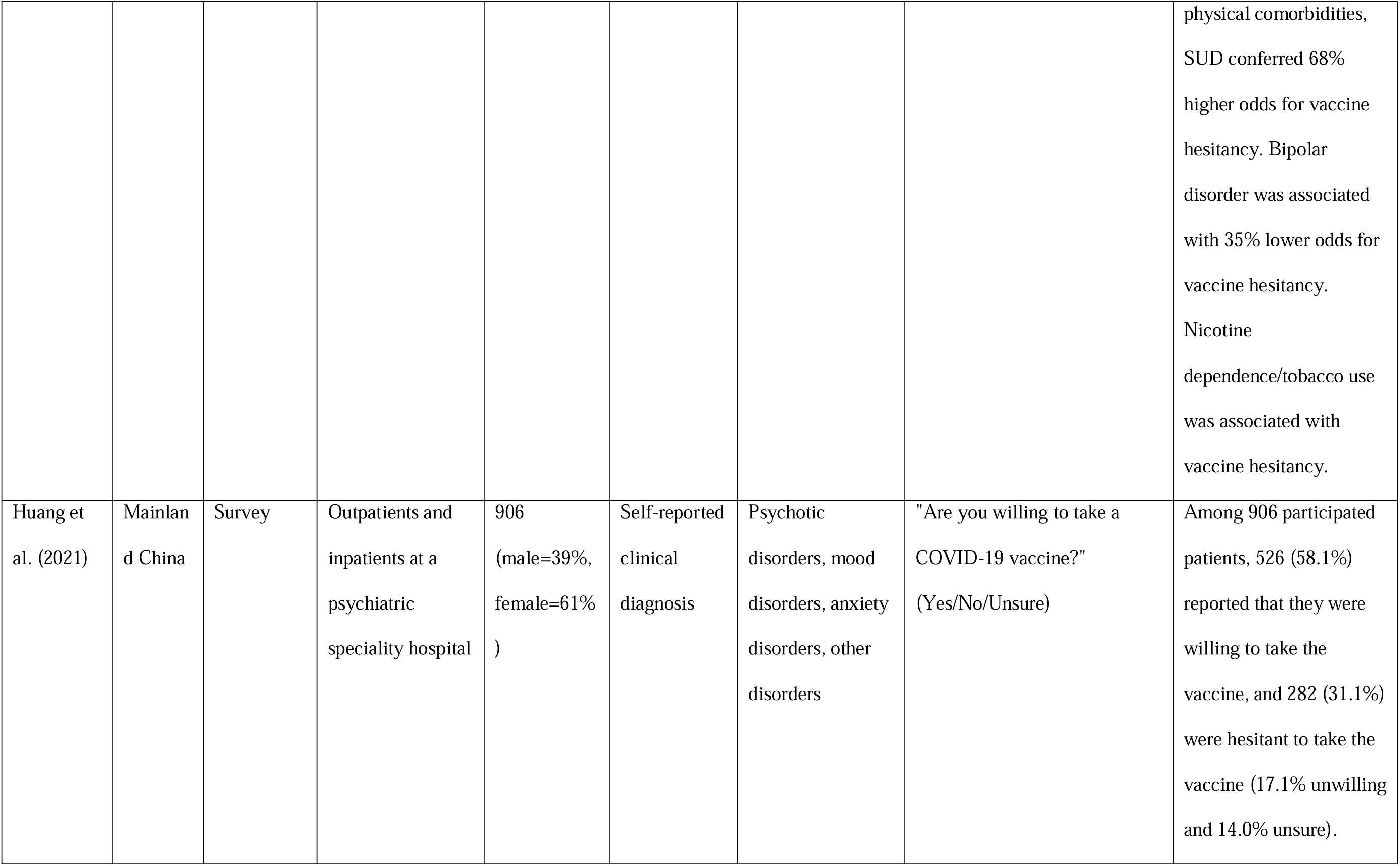
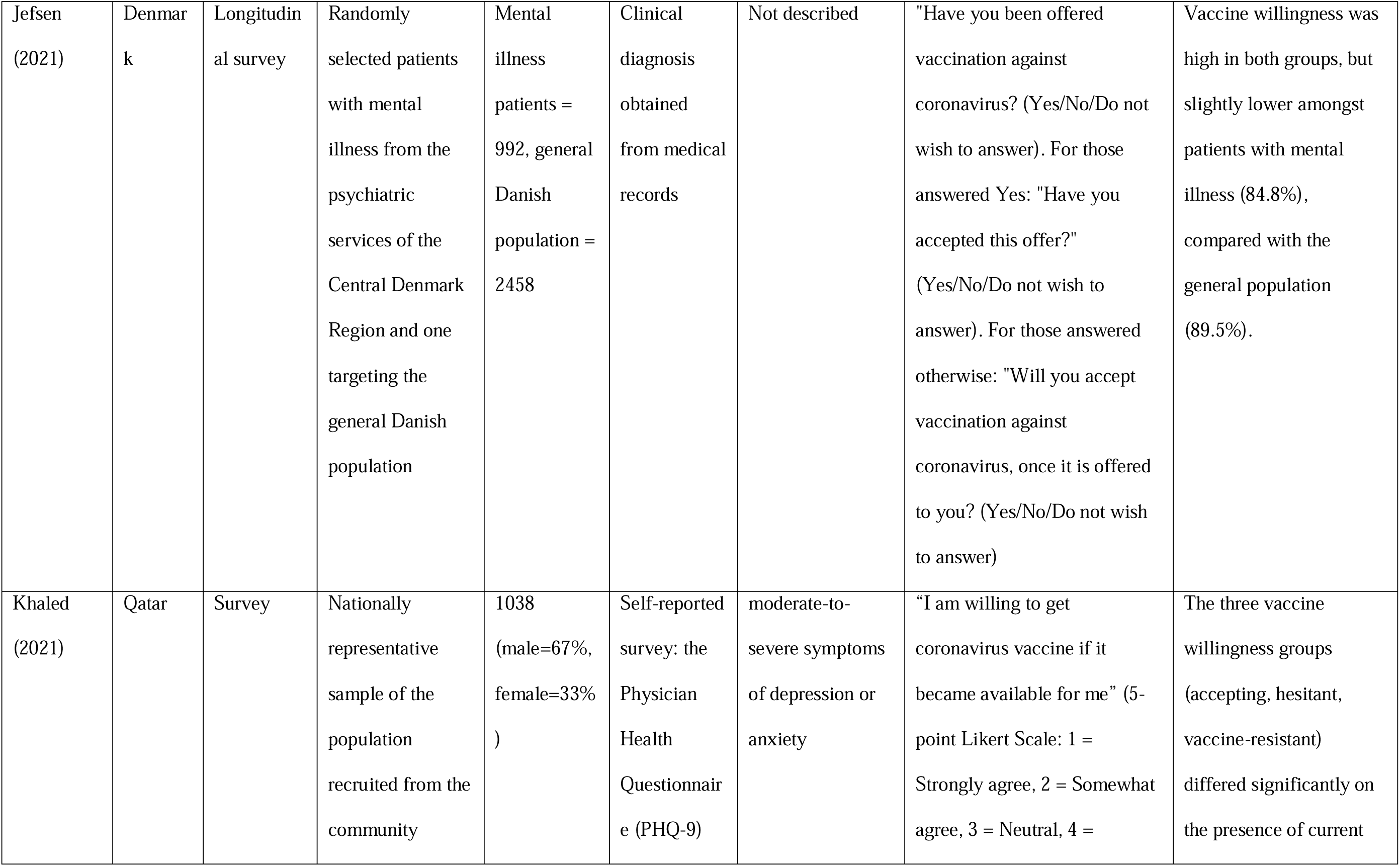
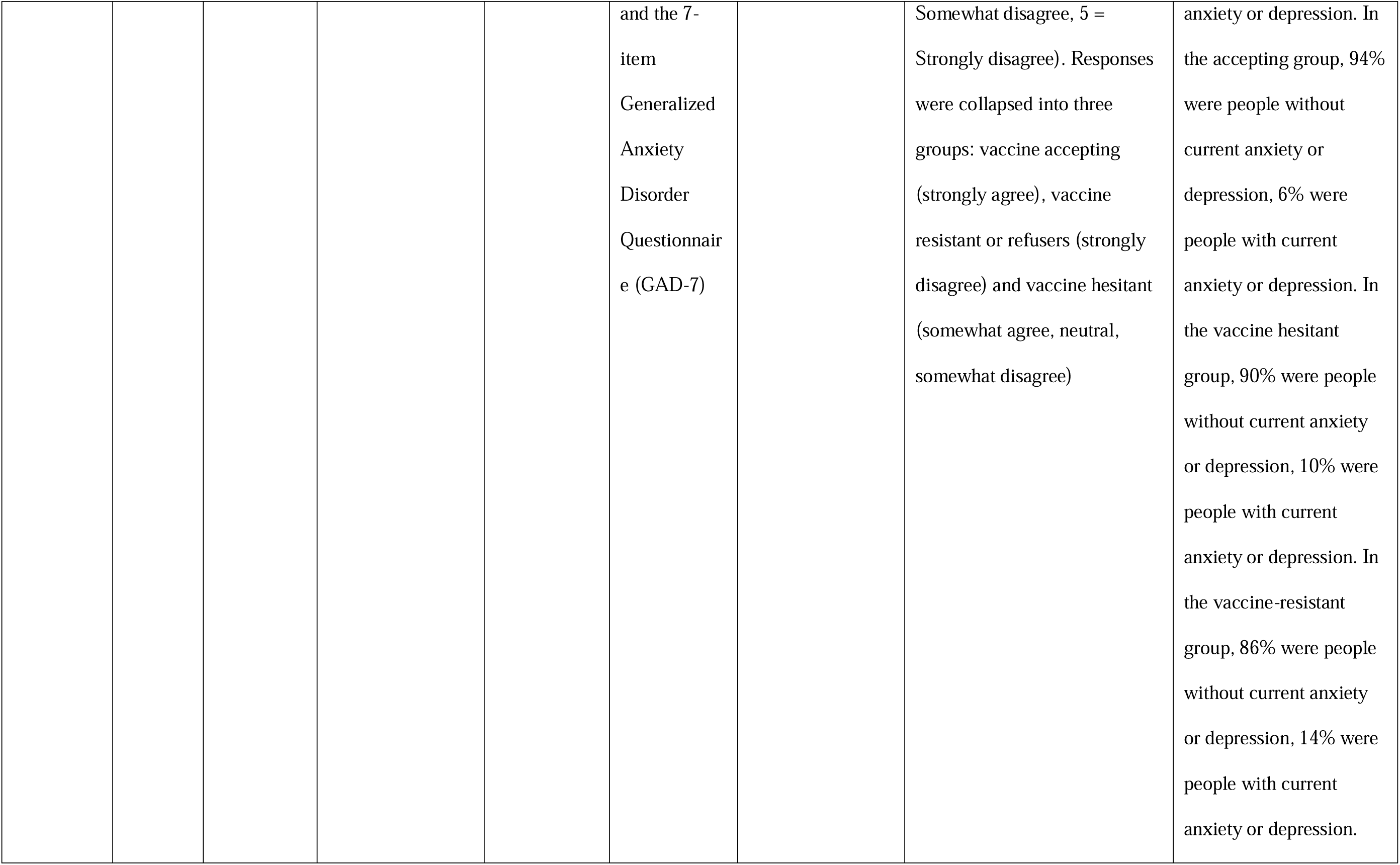
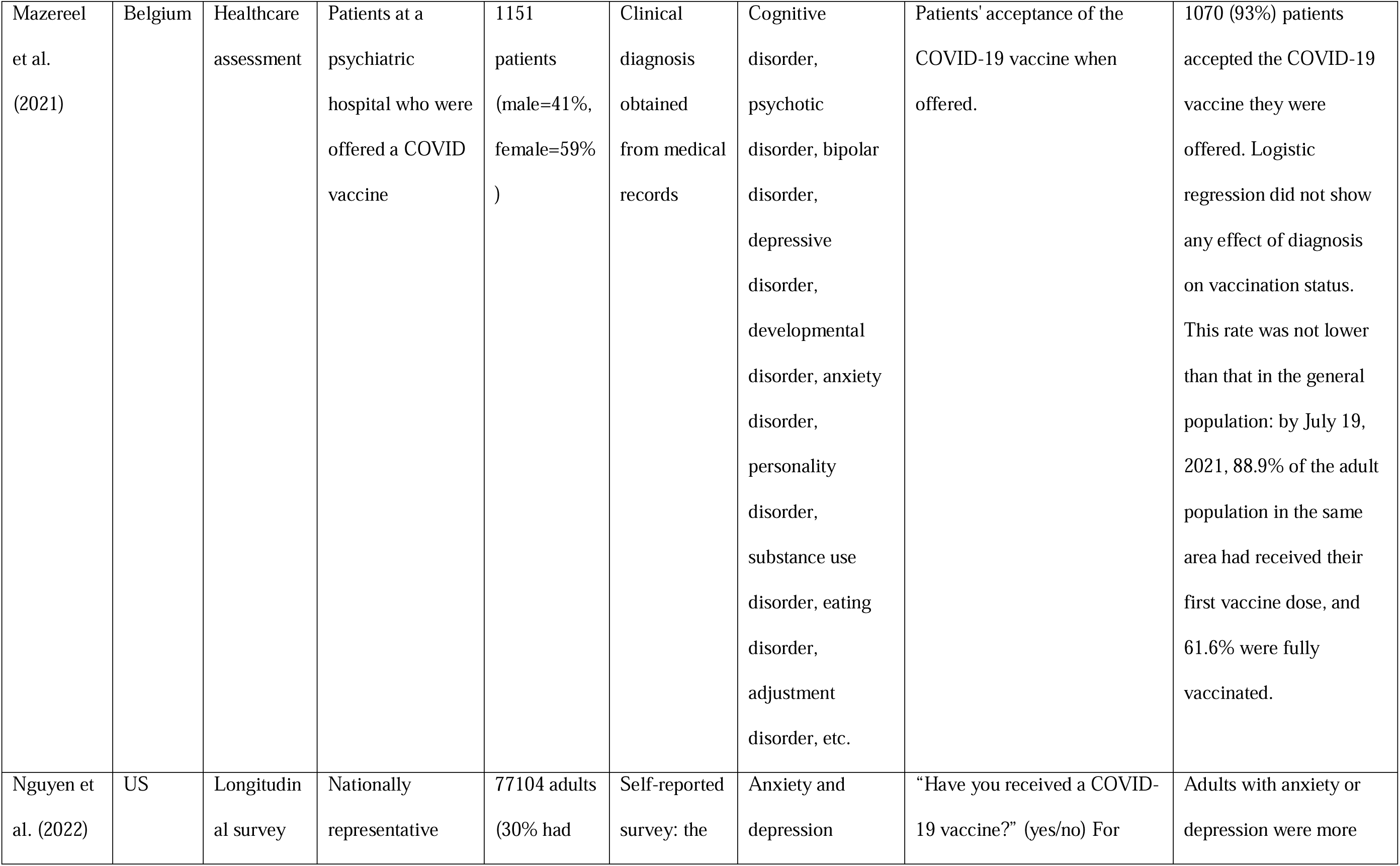
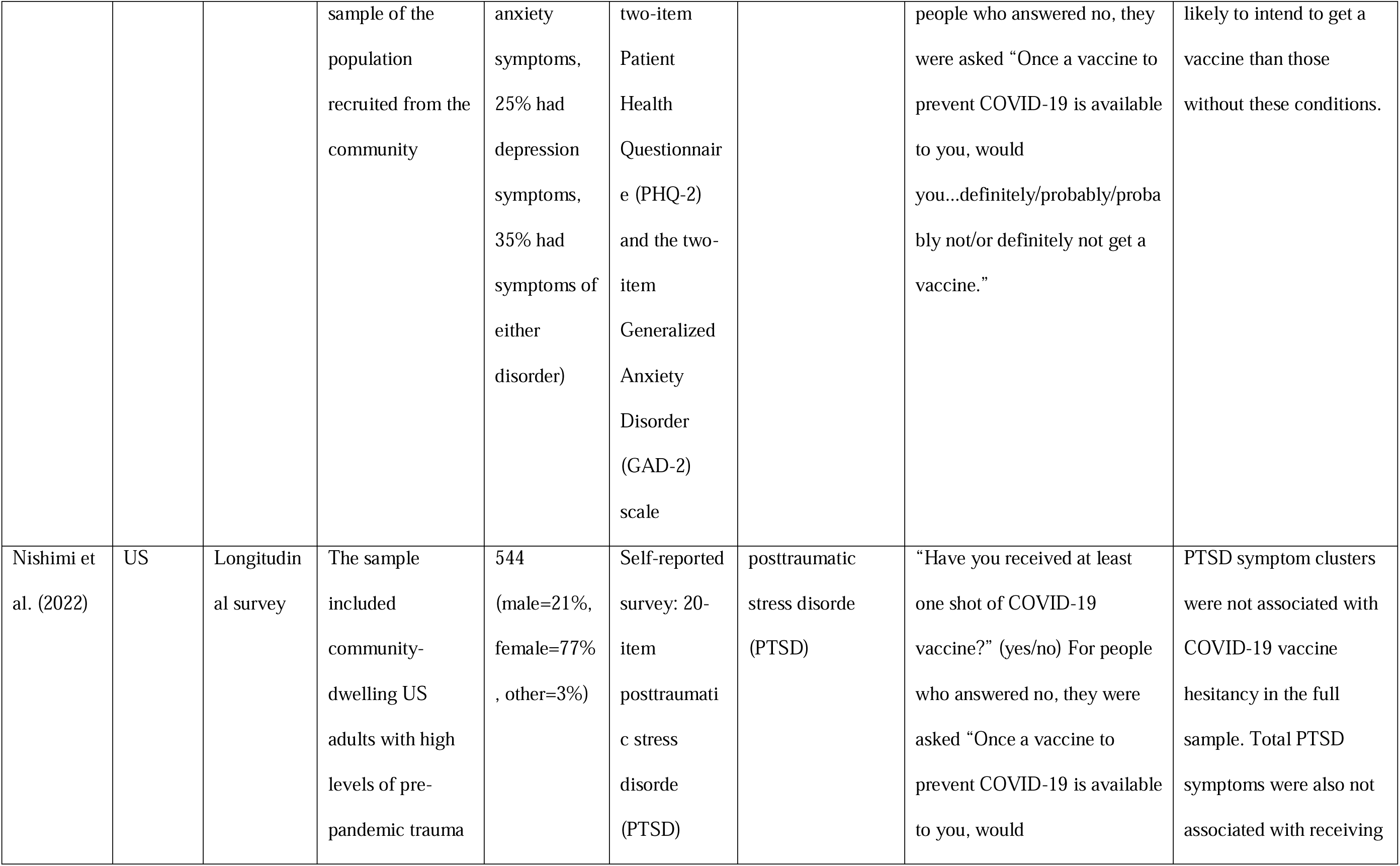
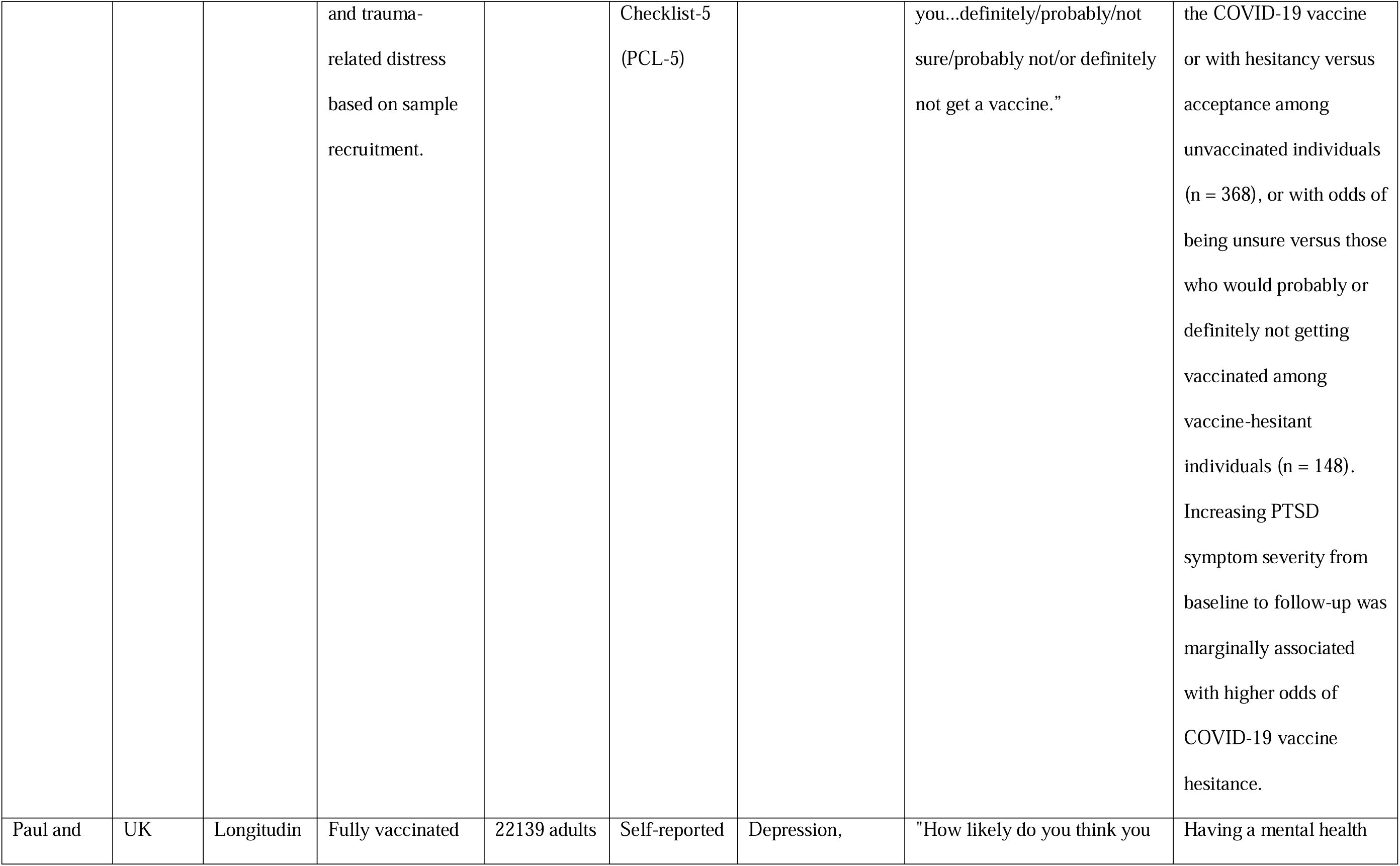
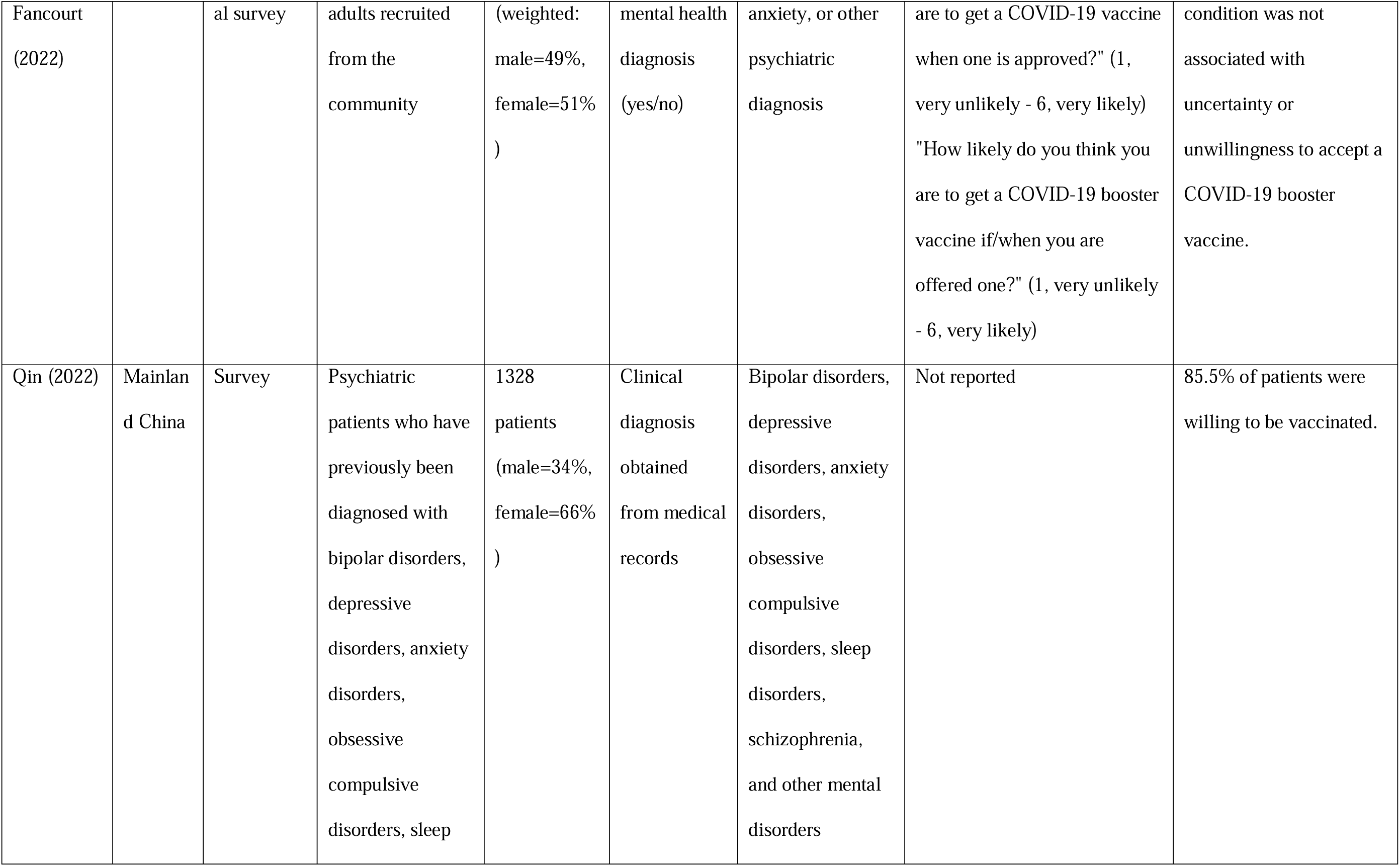
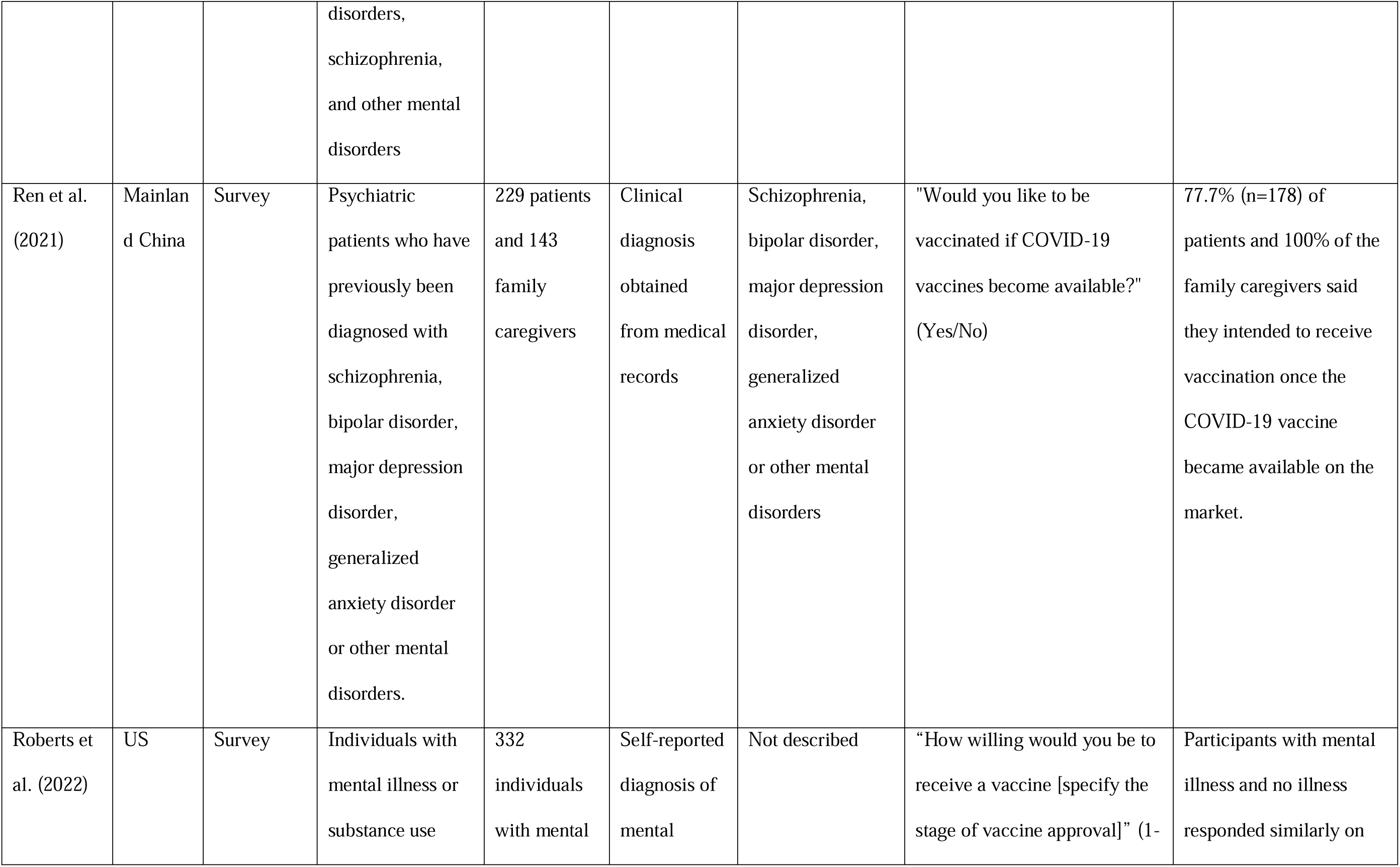
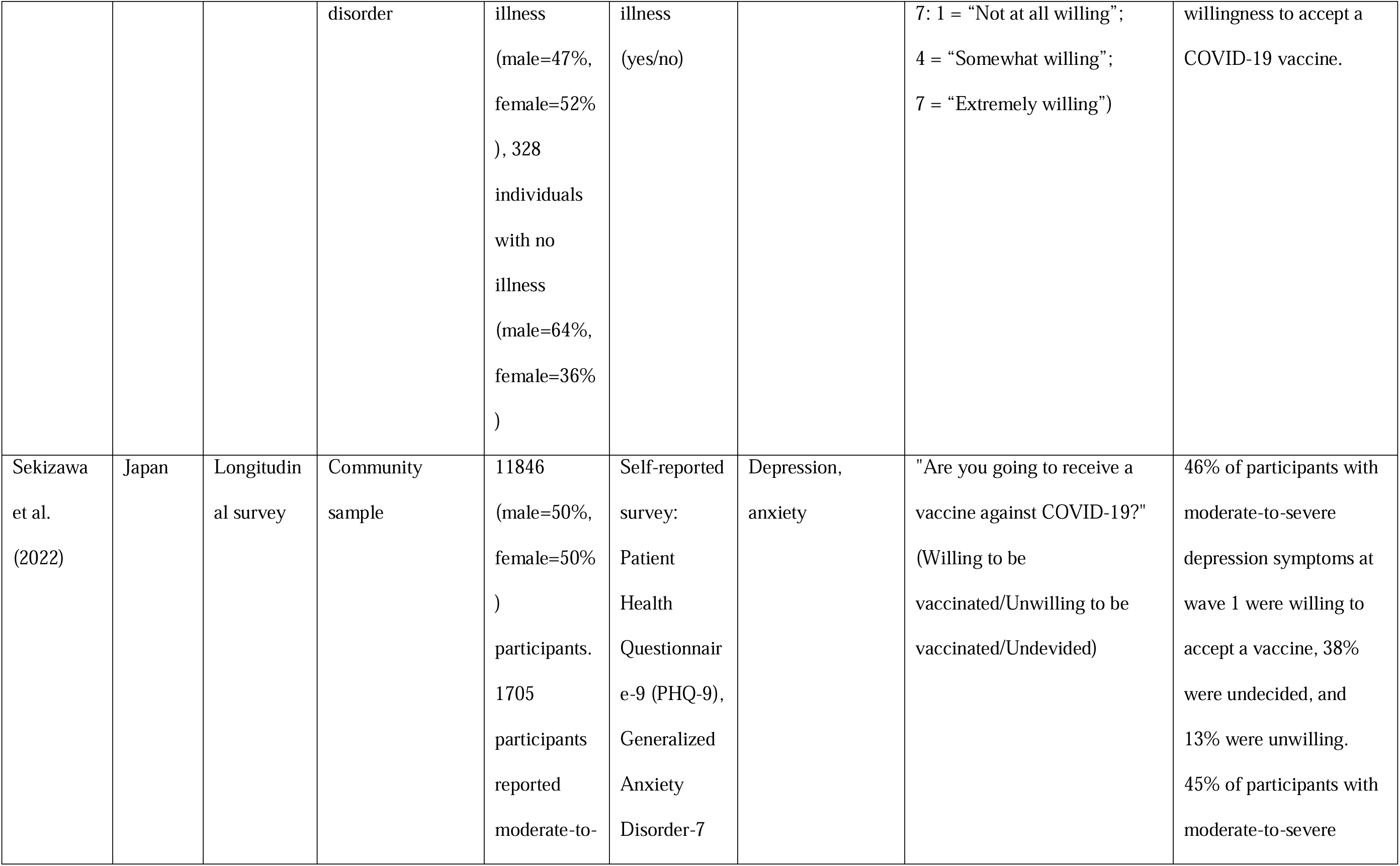
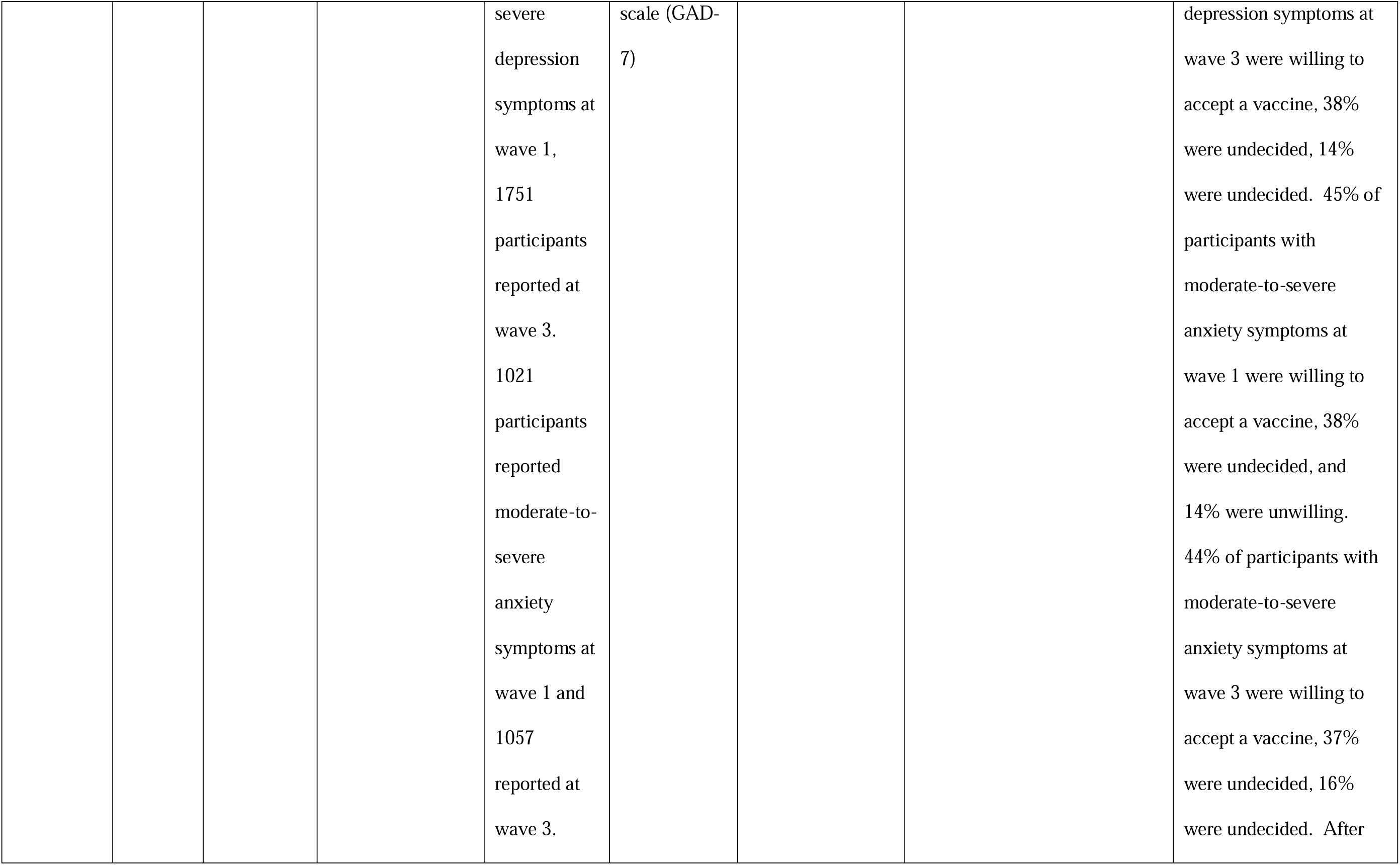
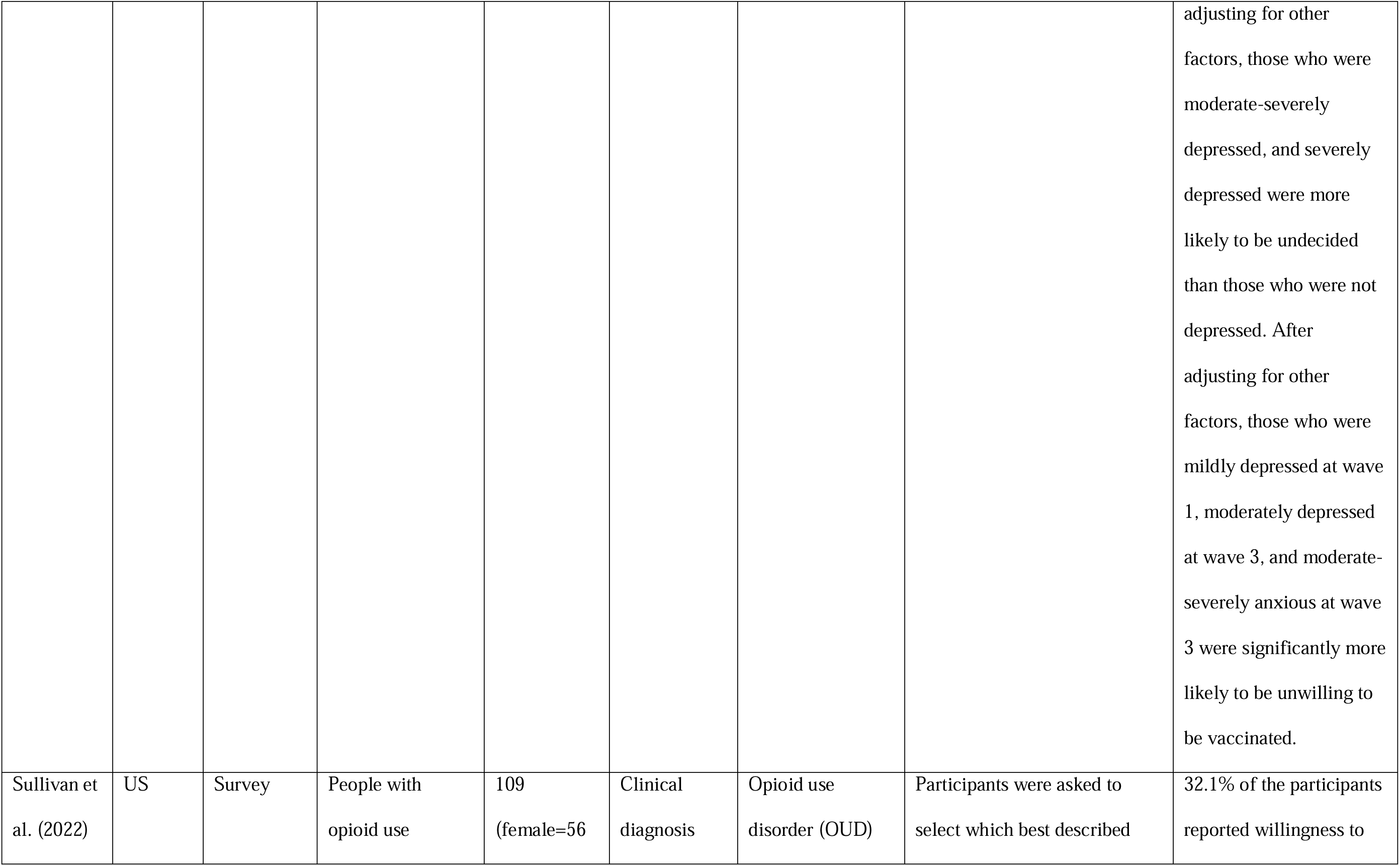
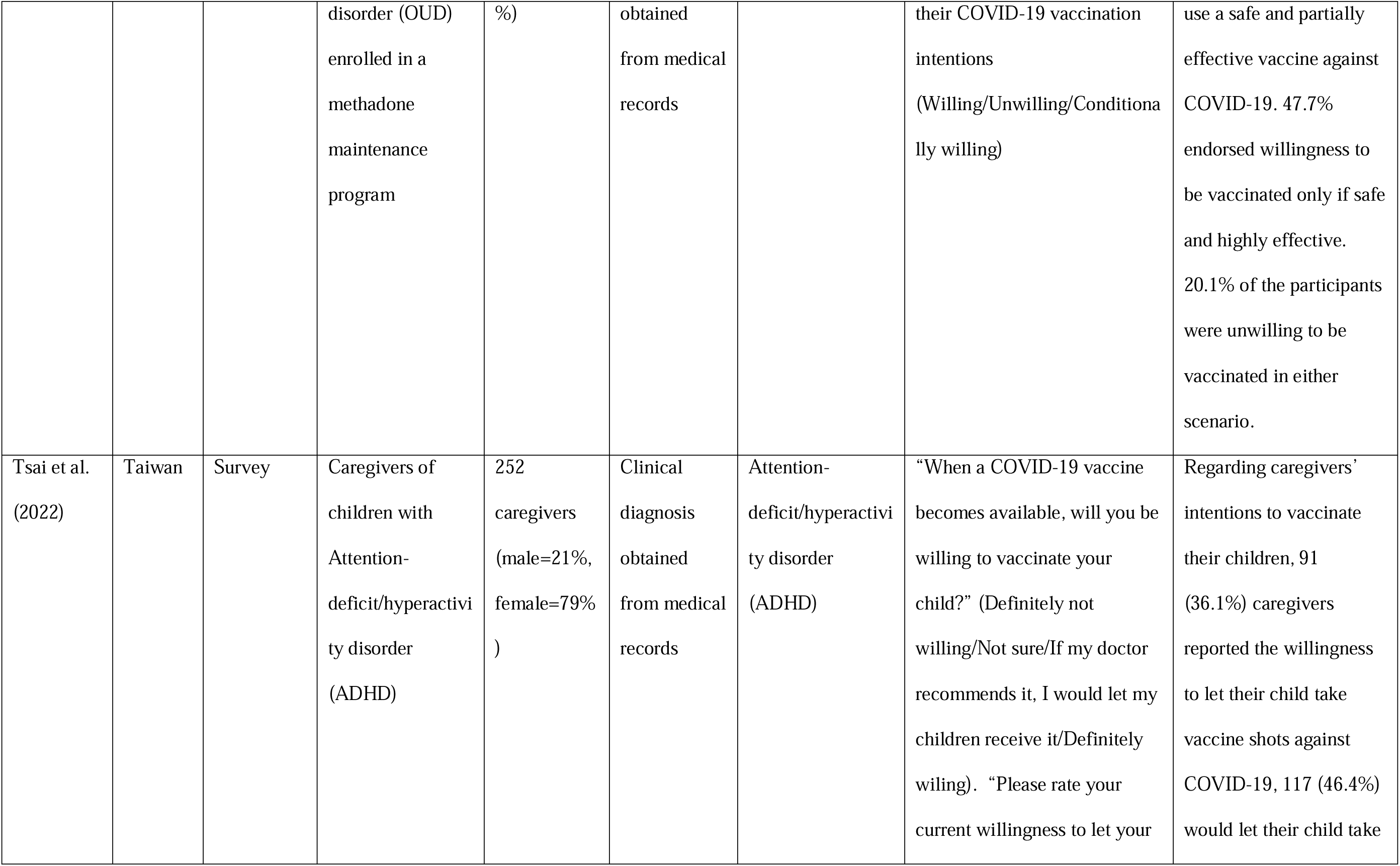
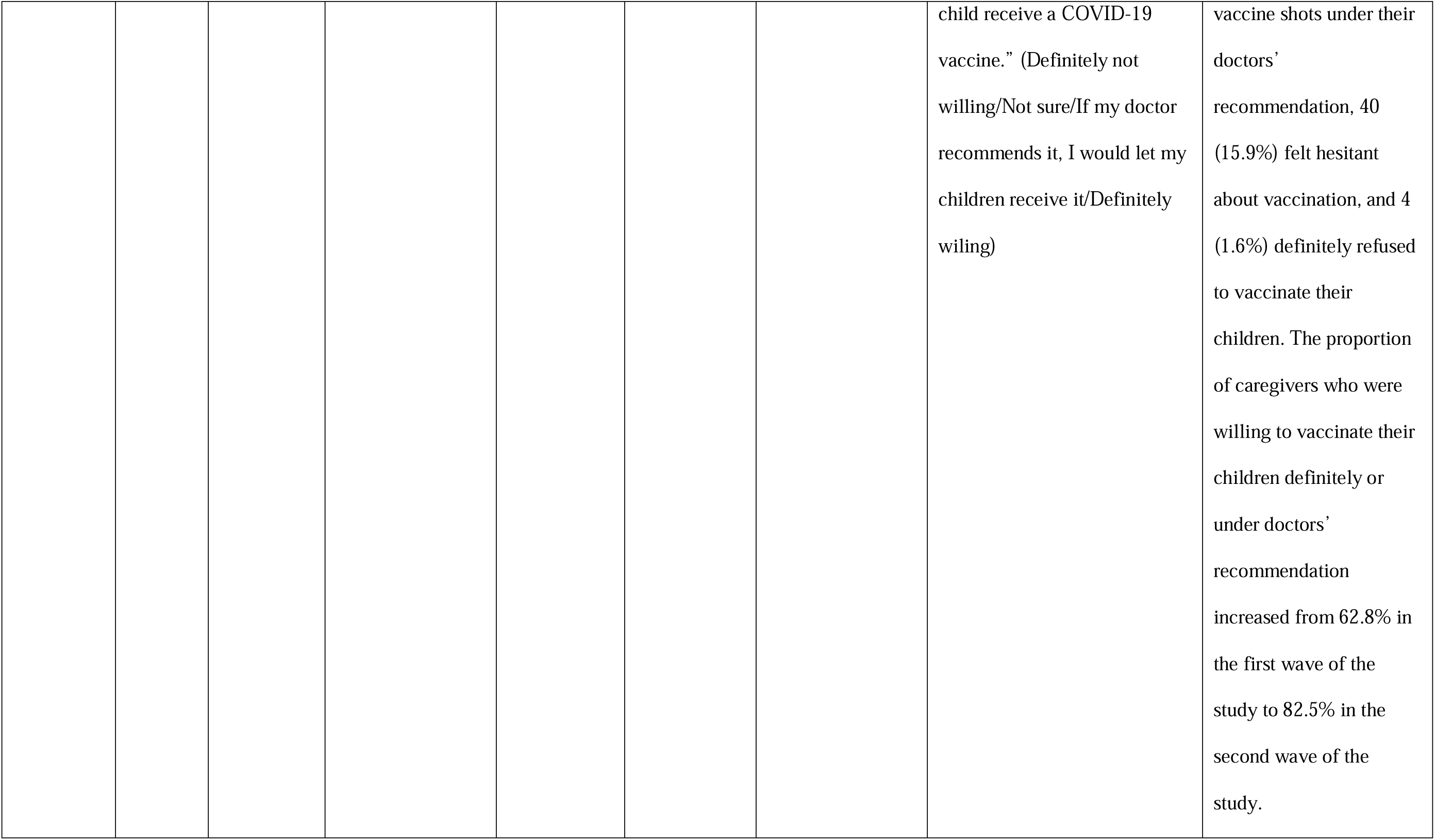
Summary of studies investigating the association between mental health conditions and COVID-19 vaccine intention.

Across the 18 studies, the intended willingness to accept a COVID-19 vaccination was between 44% and 93% among people with mental health conditions.

#### Differences between people with and without mental health conditions

When compared with people without mental health conditions, the majority of studies (i.e., 12/18) found no differences in vaccine hesitancy between people with and without mental health conditions. Specifically, 5/18 studies reported that people with mental health conditions were less willing or more hesitant to accept the COVID-19 vaccine. In general, the degree of hesitancy varied significantly between the studies with rates of hesitancy as low as 15.2% through to 53% (Abramovich et al., 2022; Dvorsky et al., 2022; Eyllon et al., 2022; Jefsen et al., 2021; Sekizawa et al., 2022). Three out of the five studies reported that the levels of hesitancy were significantly higher in patients with mental health conditions including major depressive disorder, bipolar disorder, anxiety disorder, ADHD, PTSD, substance use disorder (0.78-2.74 times, Abramovich et al., 2022; 0.65-1.68 times, Eyllon et al., 2022; 4.7%, Jefsen et al., 2021). Only 1/18 studies reported the contrary: individuals with anxiety or depression were 1.13 times (95% CI: 1.08–1.19) more likely to intend to get a vaccine than those without these conditions (Nguyen et al., 2022). Six of the 18 studies found no differences in vaccine hesitancy between people with and without mental health conditions. Specifically, one reported that having depression or anxiety symptoms was not associated with willingness to have a COVID-19 vaccine (Khaled et al., 2021). One reported that having a mental health condition was not associated with uncertainty or unwillingness to accept a COVID-19 booster vaccine (Paul & Fancourt, 2022). One reported that PTSD symptom clusters were not associated with COVID-19 vaccine hesitancy in the investigated cohort (Nishimi, Borsari, et al., 2022). The other three of the nine studies reported similar levels of willingness to accept a COVID-19 vaccine between participants with and without mental health conditions (Afifi et al., 2021; Mazereel et al., 2021; Roberts et al., 2022).

#### Differences between different mental health conditions

Three of the 18 studies reported findings in relation to patients’ different diagnosis and treatment conditions. Bai et al., (2021) did not find any significant difference in vaccine hesitancy across major depressive disorder, bipolar disorder, and schizophrenia. However, they reported that community-dwelling psychiatric patients had a higher incidence of COVID-19 vaccination hesitancy (49%) compared to hospitalised patients (31%). Mazereel et al., (2021) also reported no effect of different psychiatric diagnosis on vaccine acceptance. Eyllon et al., (2022) reported that, after adjusting for other sociodemographic characteristics and physical comorbidities, patients with substance use disorders had 68% higher odds for vaccine hesitancy while bipolar disorder was associated with 35% lower odds for vaccine hesitancy.

Overall, these findings suggest that most studies fail to find evidence of significantly greater hesitancy in people with mental health conditions, with only five studies reporting greater hesitancy among people with mental health conditions. The evidence on the association between different mental health diagnosis and vaccine intentions is limited and, therefore, not conclusive. The available evidence suggests that patients with substance use disorder might have higher risks of COVID-19 vaccine hesitancy, compared with other mental health conditions.

### Mental health conditions and COVID-19 vaccine uptake

Seventeen of the 33 reviewed studies reported on the association between mental health conditions and the uptake of the COVID-19 vaccine or booster vaccine (Table 2, Balut et al., 2021; Curtis, Inglesby, MacKenna, et al., 2022; Curtis, Inglesby, Morton, et al., 2022; Gibbon et al., 2021; Hassan et al., 2022; Huang et al., 2021; Mazereel et al., 2021; Moeller et al., 2021; Murphy et al., 2022; Nguyen et al., 2022; Nilsson et al., 2022; Qin et al., 2022; Raffard et al., 2022; Sekizawa et al., 2022; Shkalim Zemer et al., 2022; Tzur Bitan et al., 2022; Uvais, 2022). Sizes of the studies ranged from N=62 to 57.9 million. Nine of the 17 studies measured COVID-19 vaccine uptake using participants’ health or medical records (Balut et al., 2021; Curtis, Inglesby, MacKenna, et al., 2022; Curtis, Inglesby, Morton, et al., 2022; Hassan et al., 2022; Moeller et al., 2021; Murphy et al., 2022; Nilsson et al., 2022; Shkalim Zemer et al., 2022; Tzur Bitan et al., 2022), five of the 17 studies asked participants to self-report their vaccine status (Huang et al., 2021; Nguyen et al., 2022; Raffard et al., 2022; Sekizawa et al., 2022; Uvais, 2022), and two of the 17 studies recorded the number of patients in hospital accepting the COVID-19 vaccine when offered (Gibbon et al., 2021; Mazereel et al., 2021). One of the 17 studies did not clearly describe how vaccine uptake was measured (Qin et al., 2022). 13 of the 17 studies reported on people with current mental health conditions (Balut et al., 2021; Gibbon et al., 2021; Huang et al., 2021; Mazereel et al., 2021; Moeller et al., 2021; Murphy et al., 2022; Nguyen et al., 2022; Qin et al., 2022; Raffard et al., 2022; Sekizawa et al., 2022; Shkalim Zemer et al., 2022; Tzur Bitan et al., 2022; Uvais, 2022). Four of the 17 studies reported on people with either current or history of mental health conditions, but no distinction was made between current and history of mental health conditions (Curtis, Inglesby, MacKenna, et al., 2022; Curtis, Inglesby, Morton, et al., 2022; Hassan et al., 2022; Nilsson et al., 2022). Therefore, we did not distinguish between current and history of mental health conditions in the following reporting.

**Table 2.**
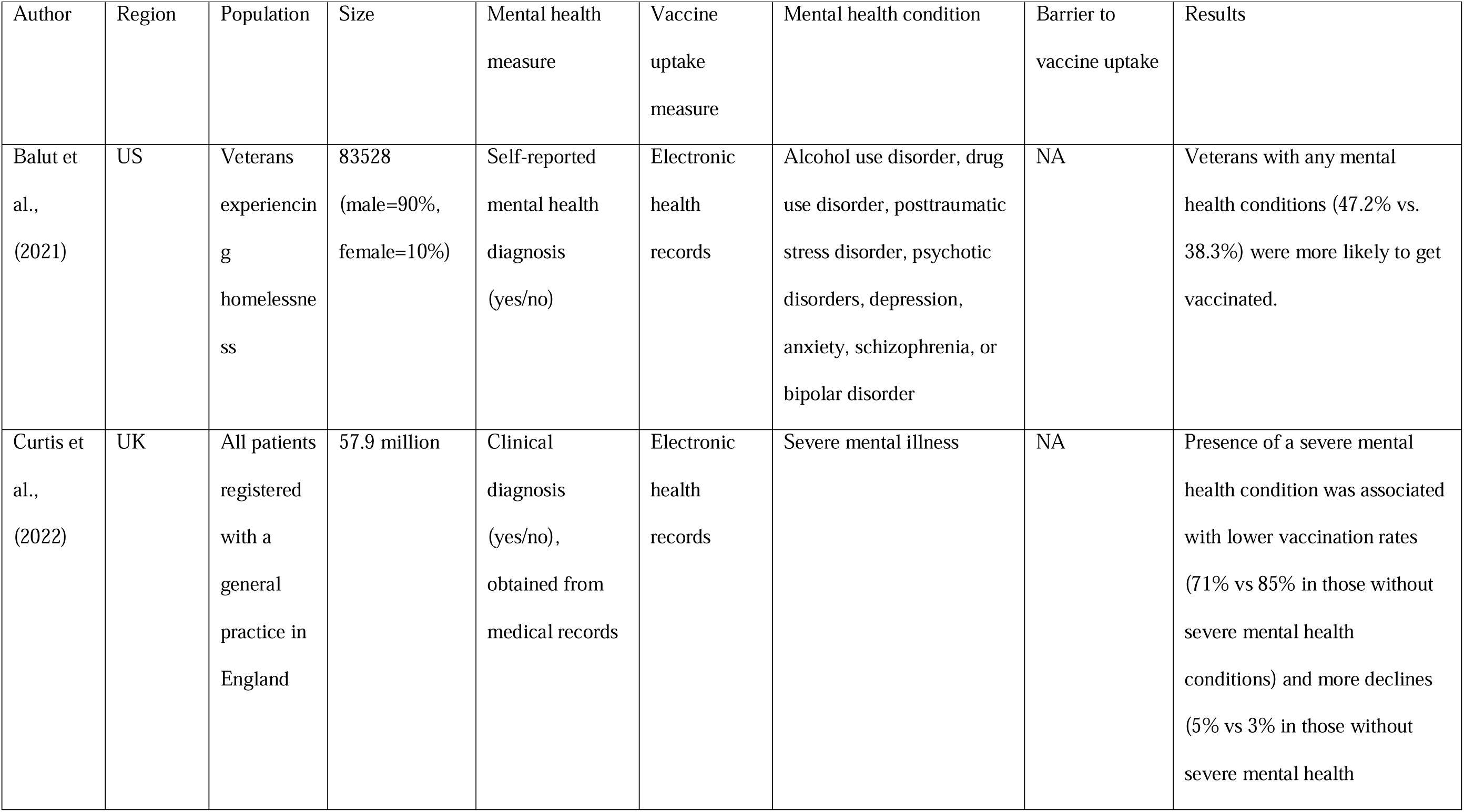
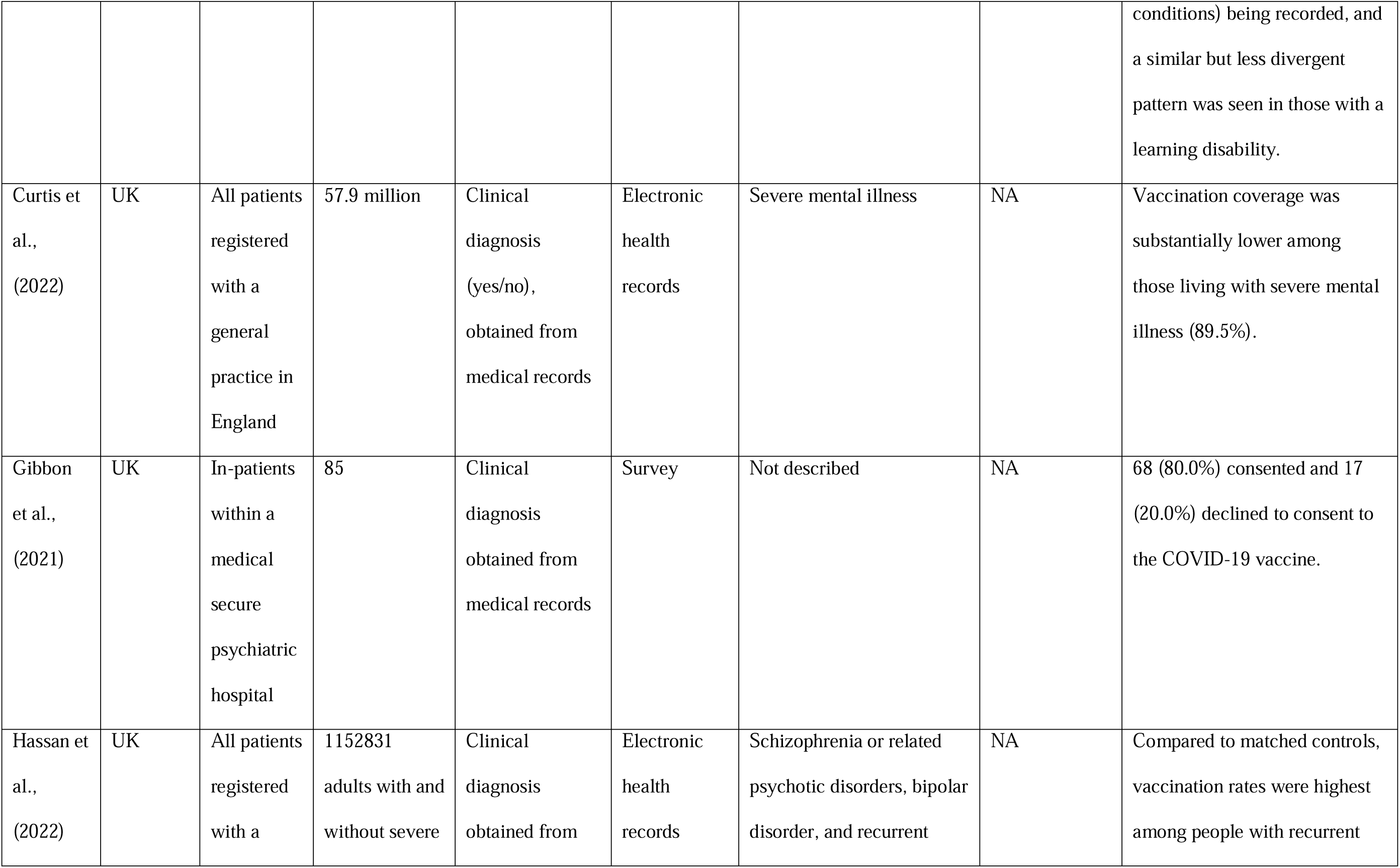
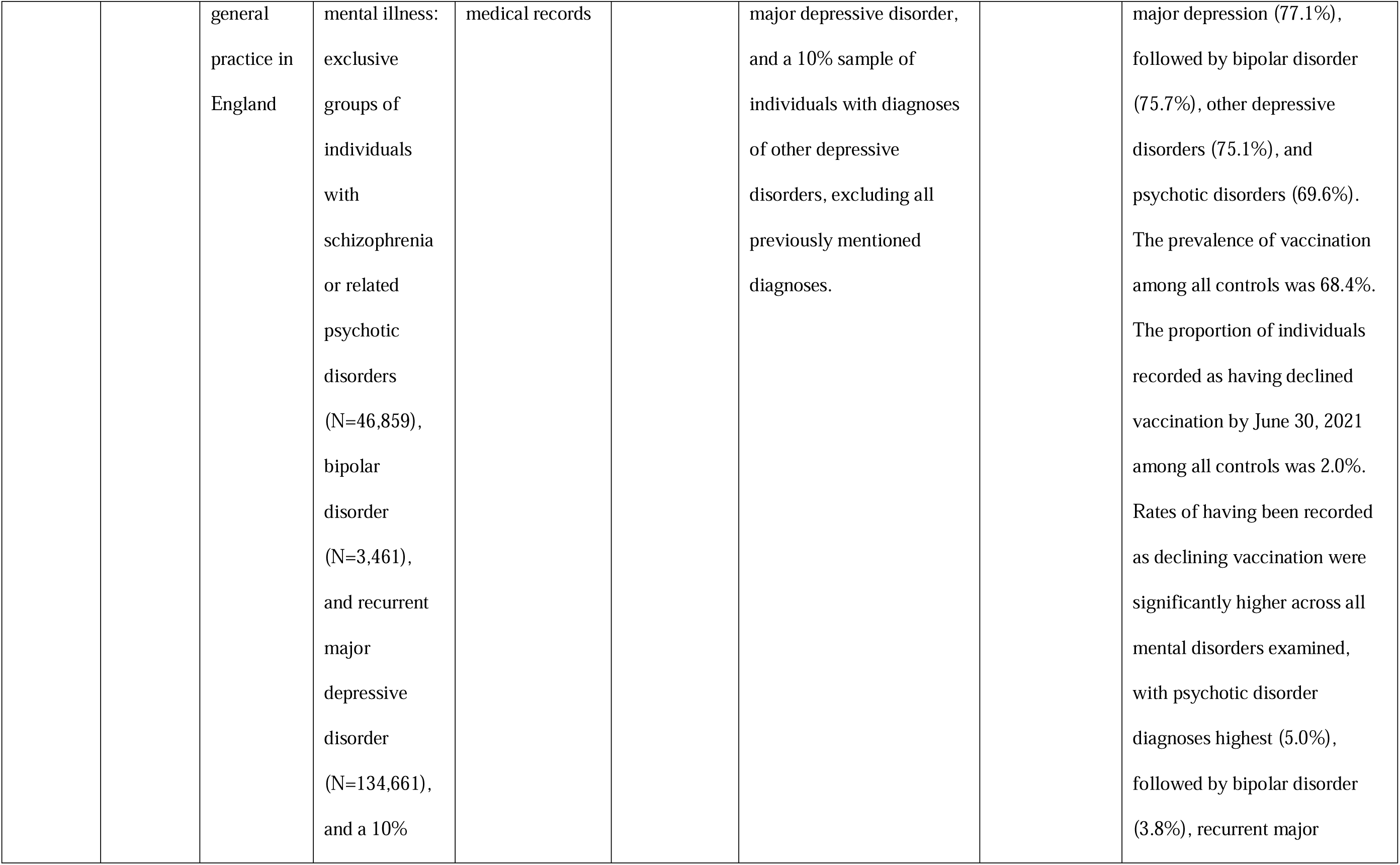
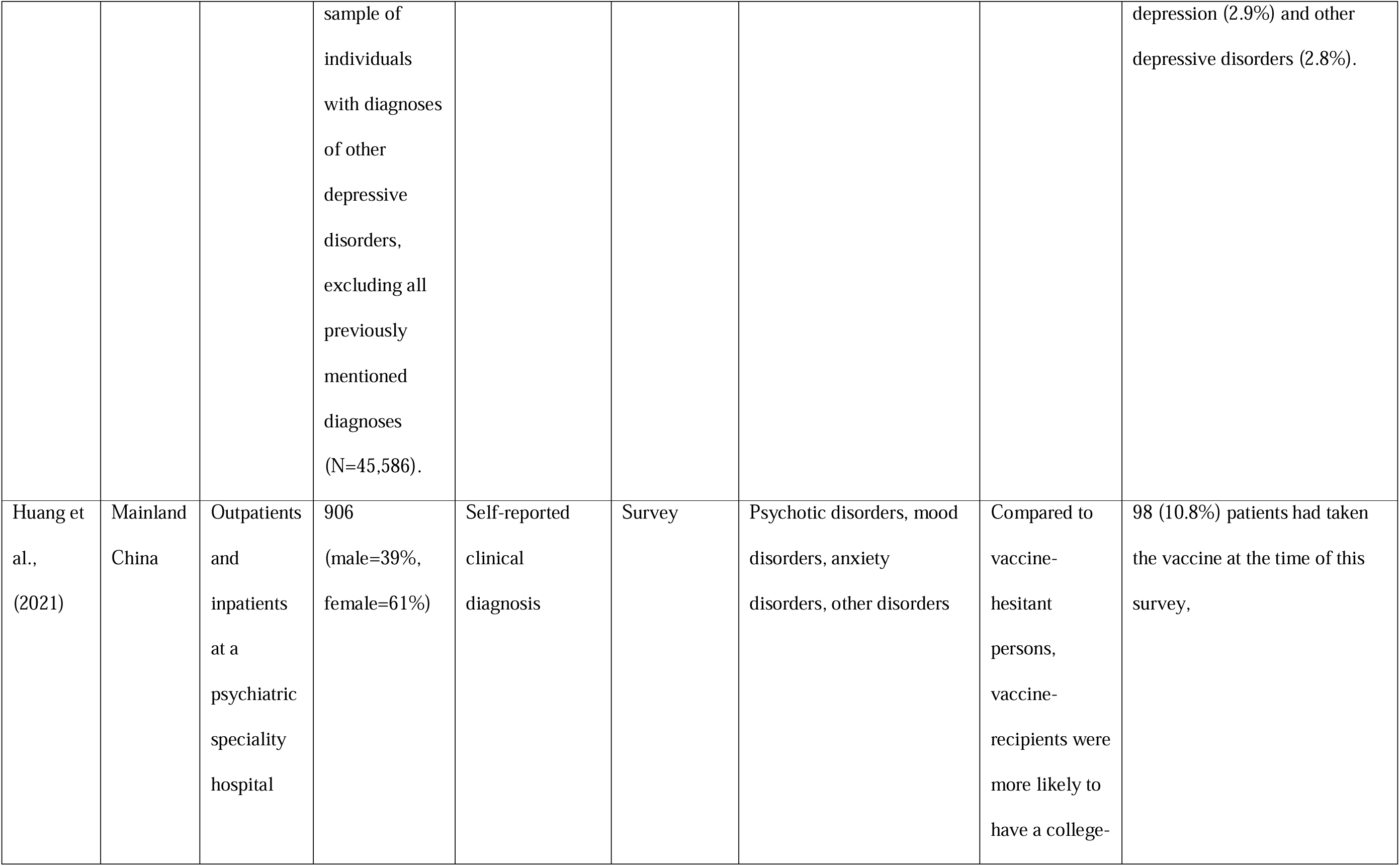
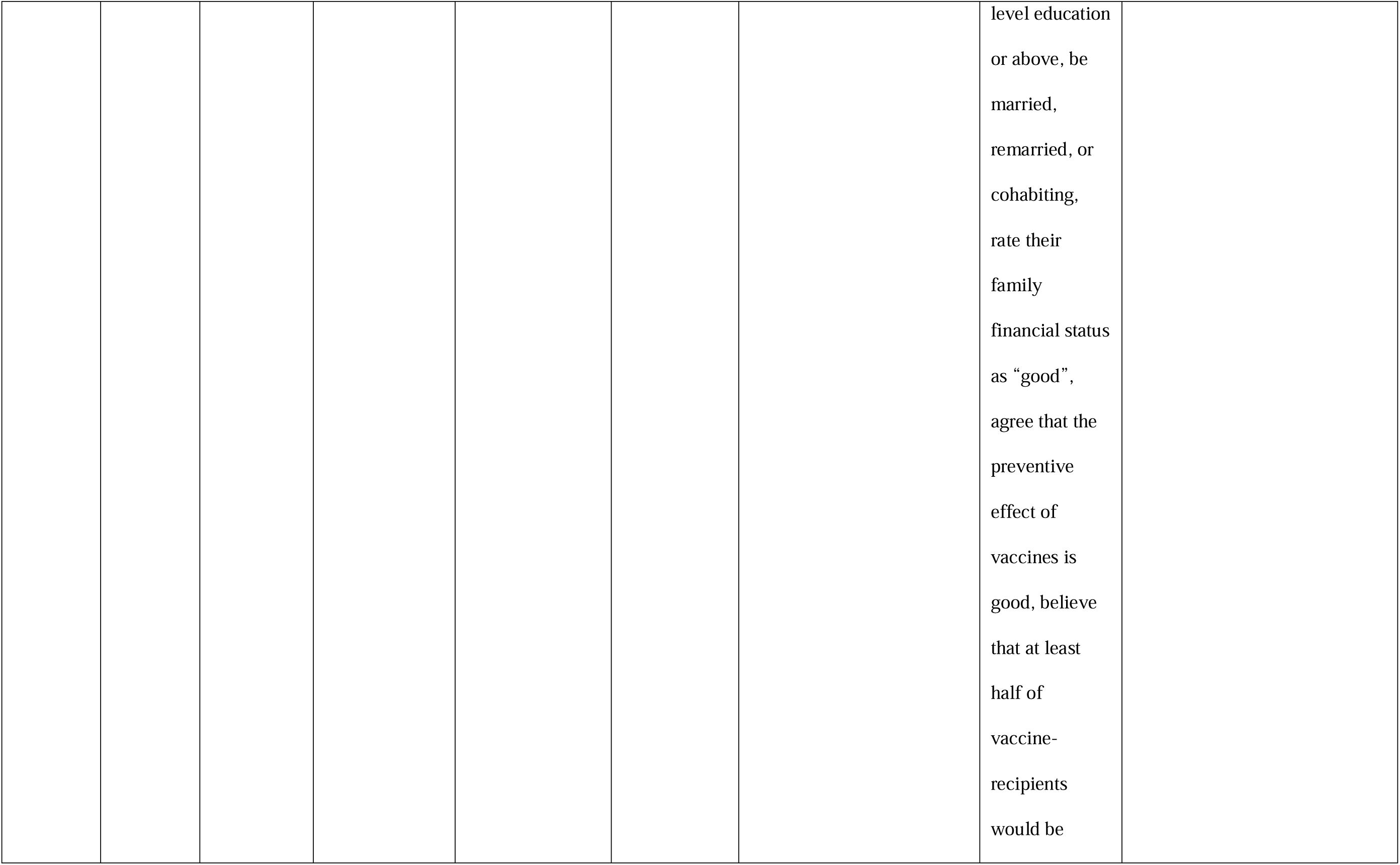
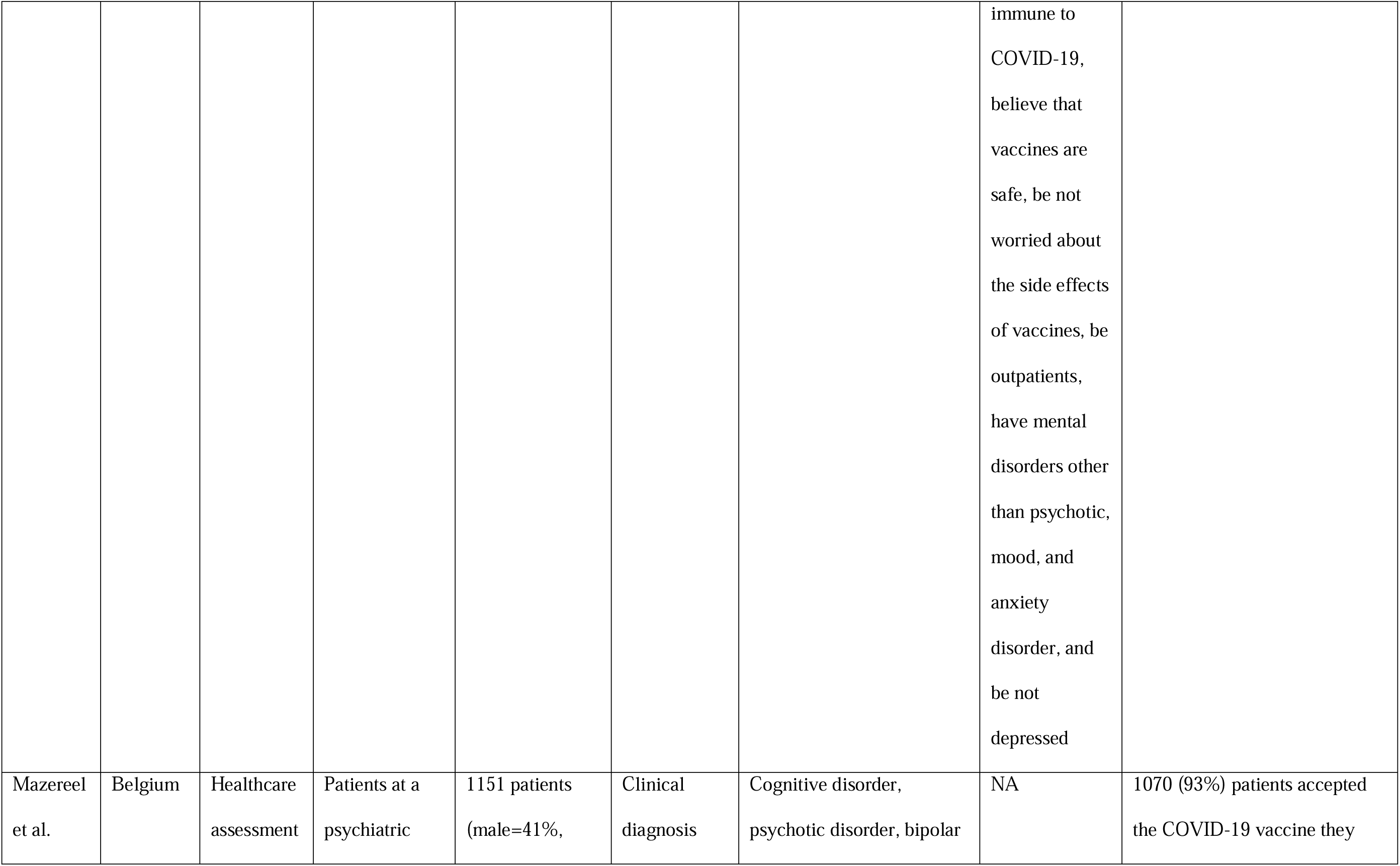
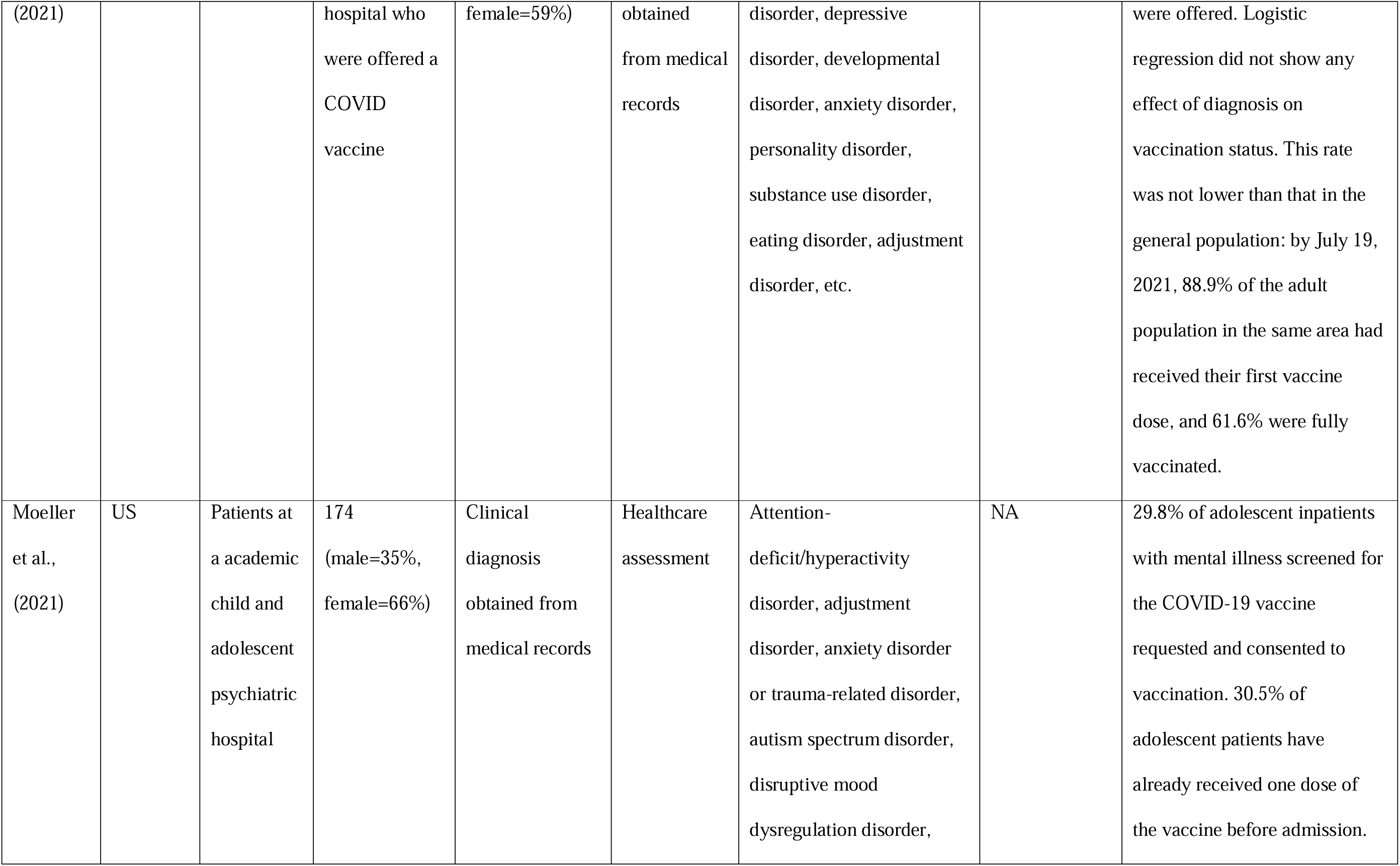
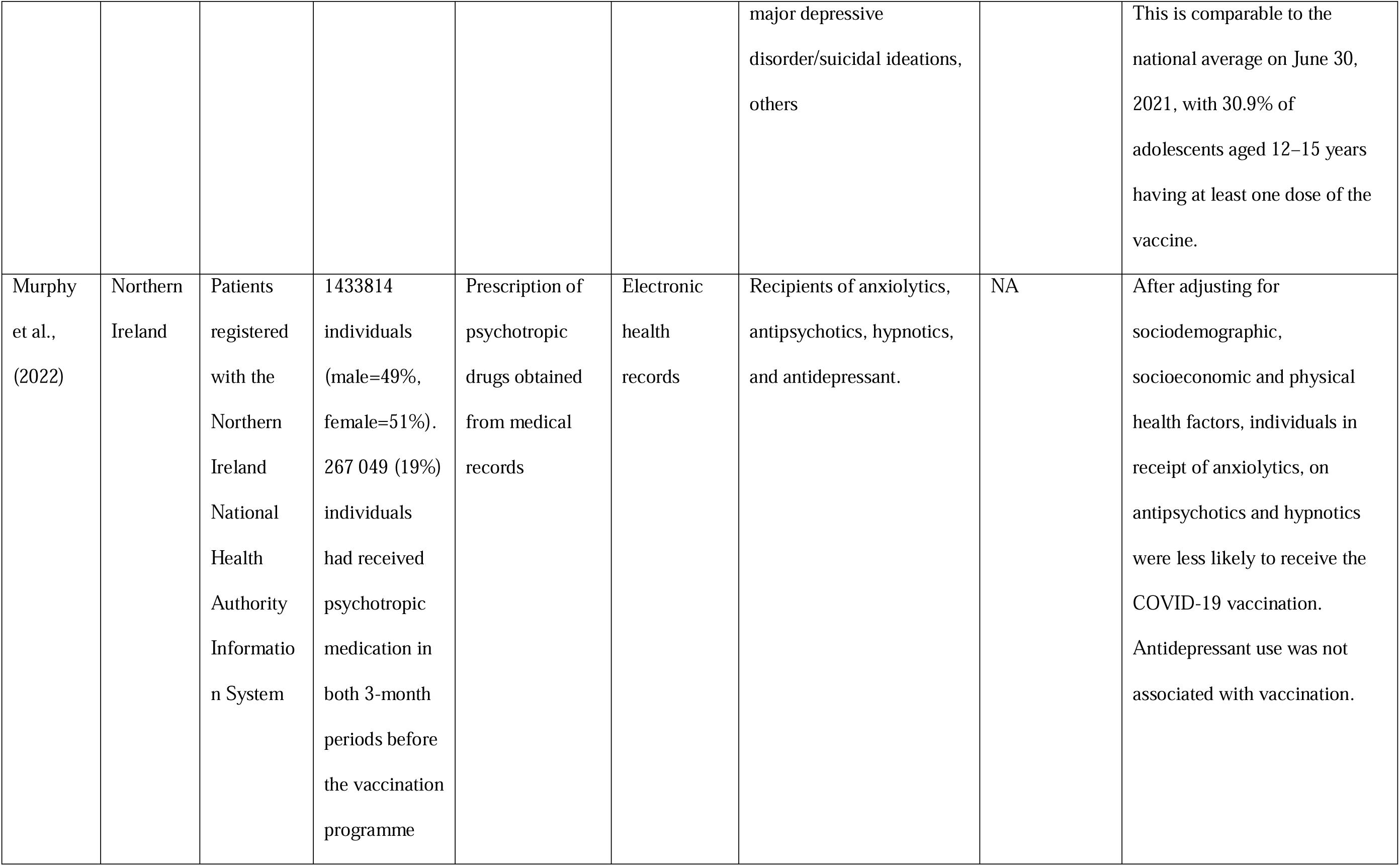
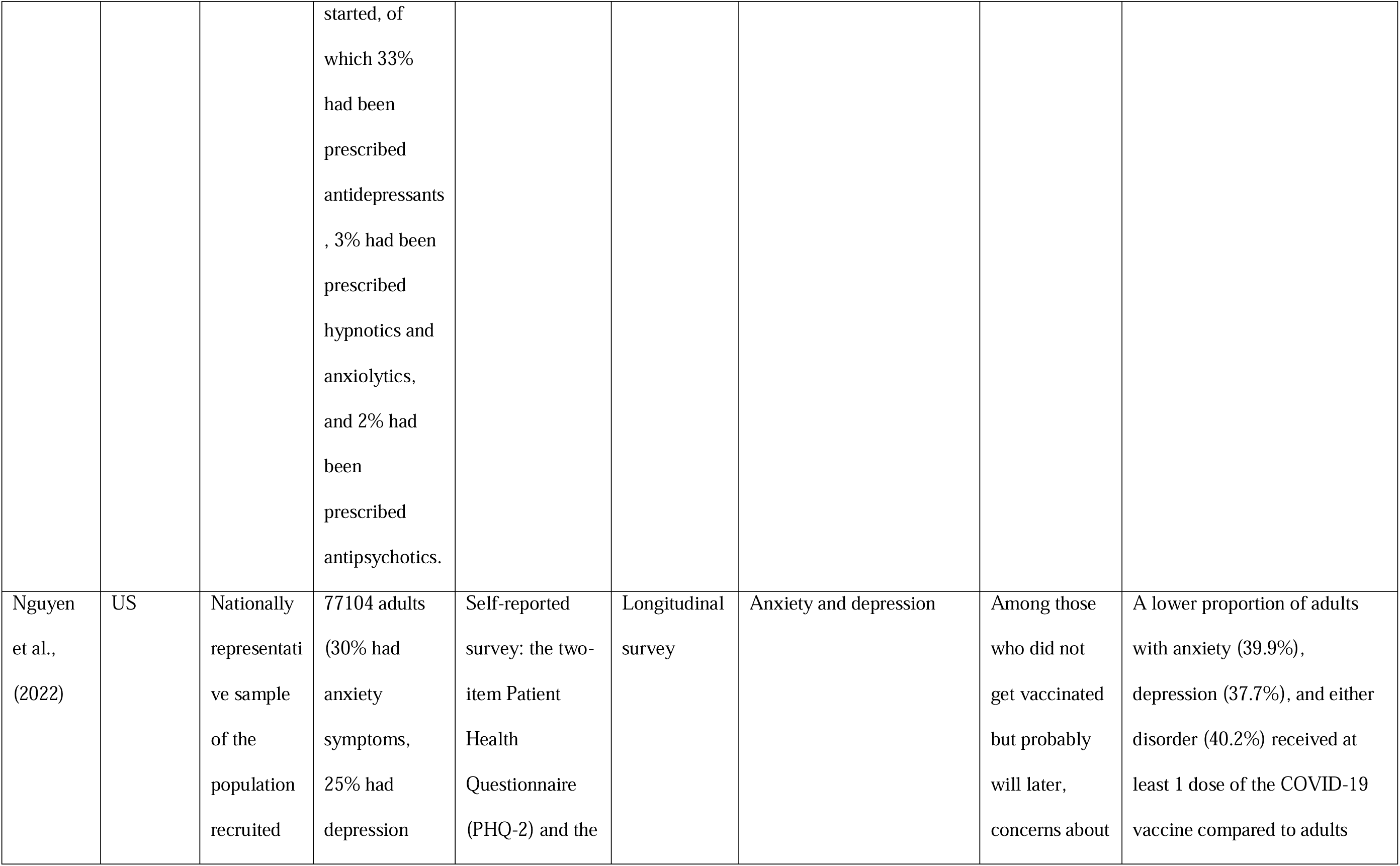
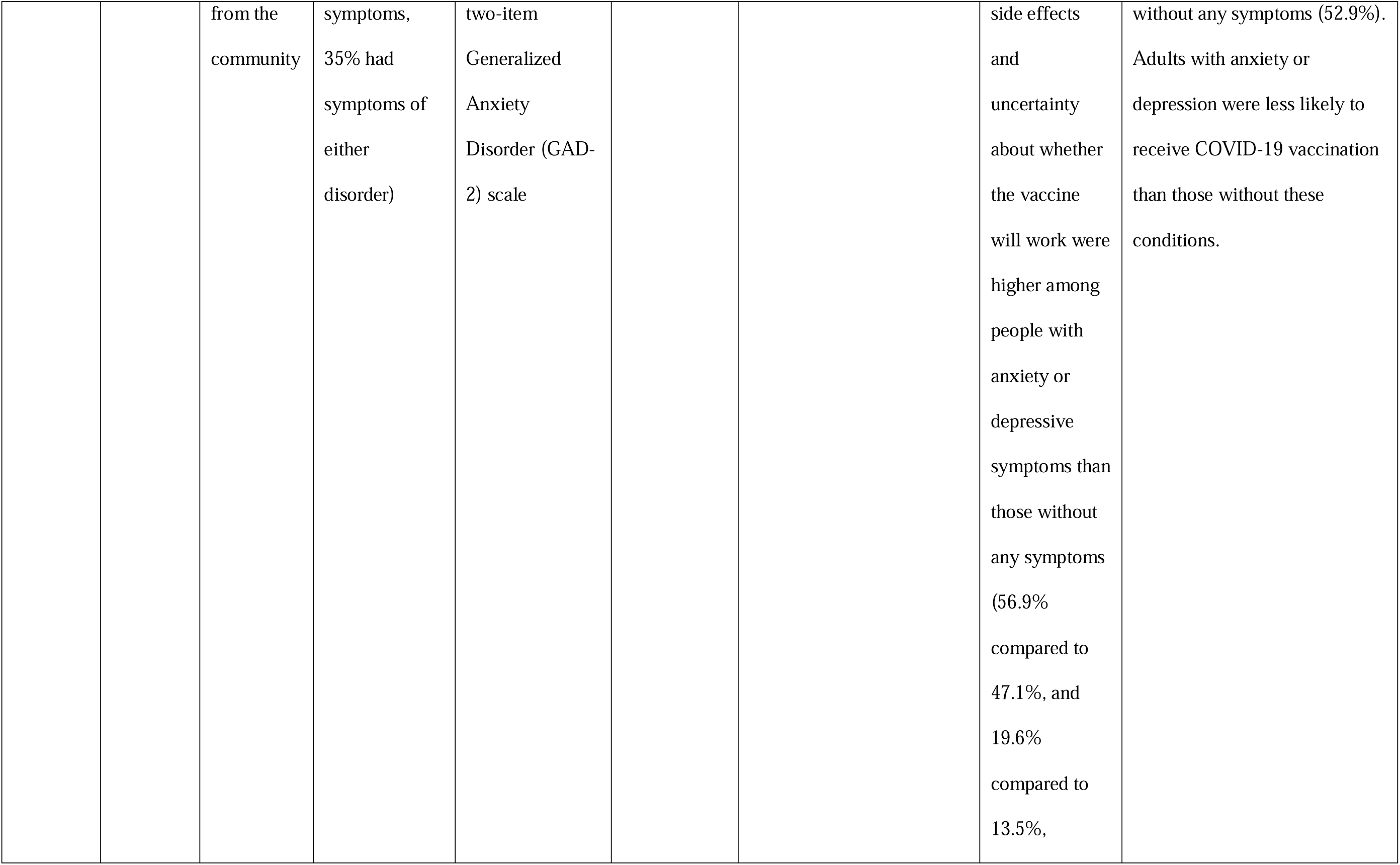
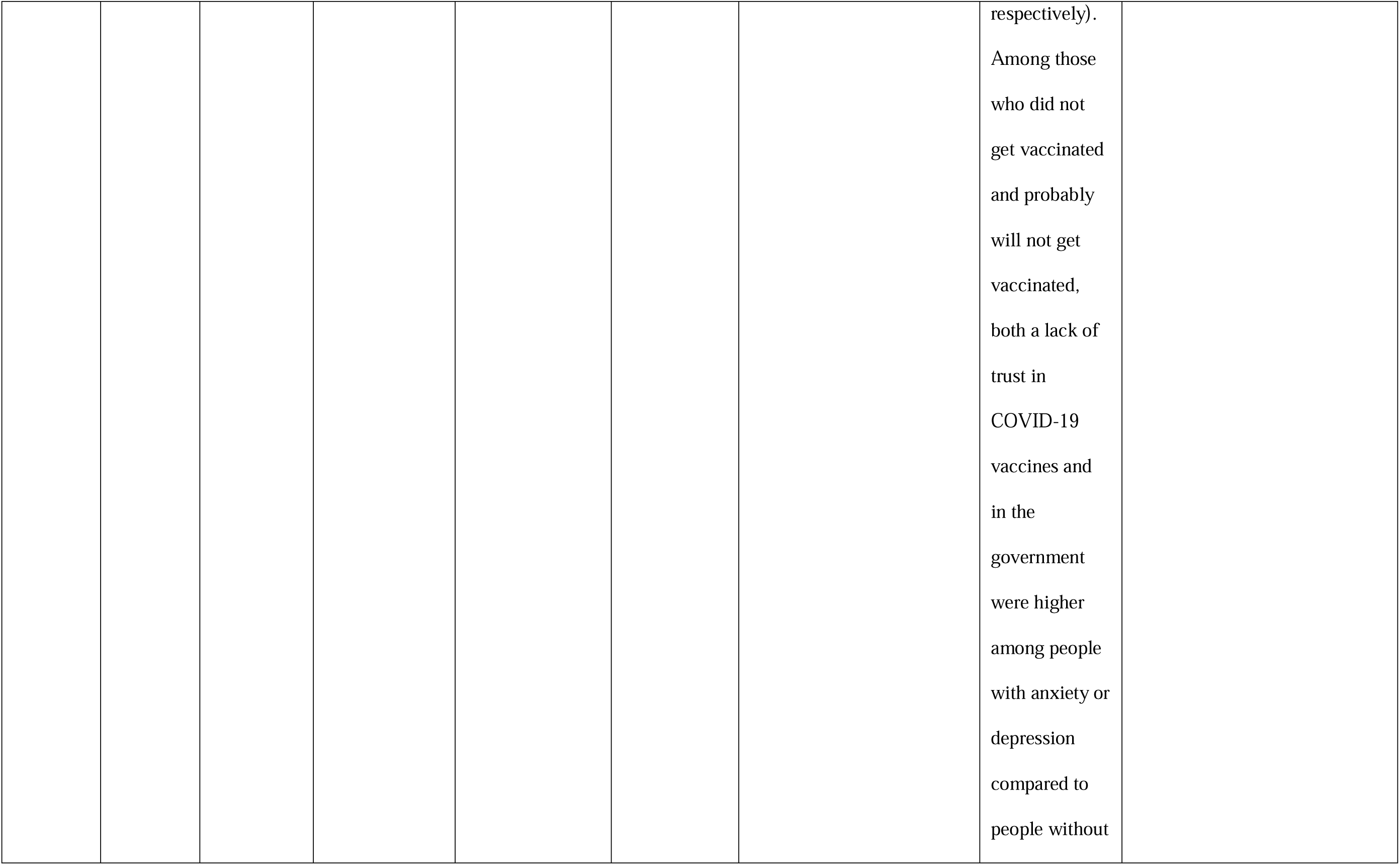
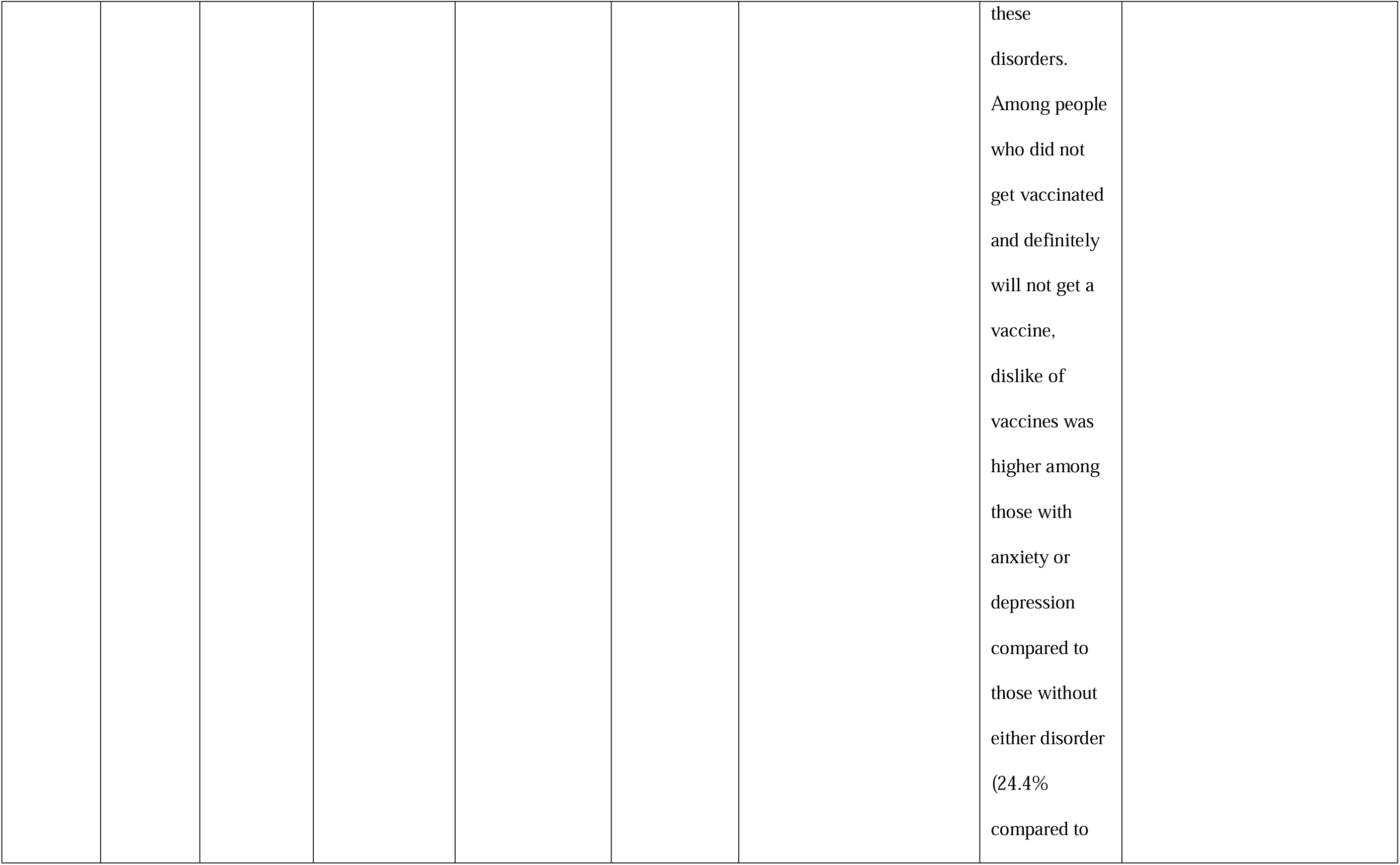
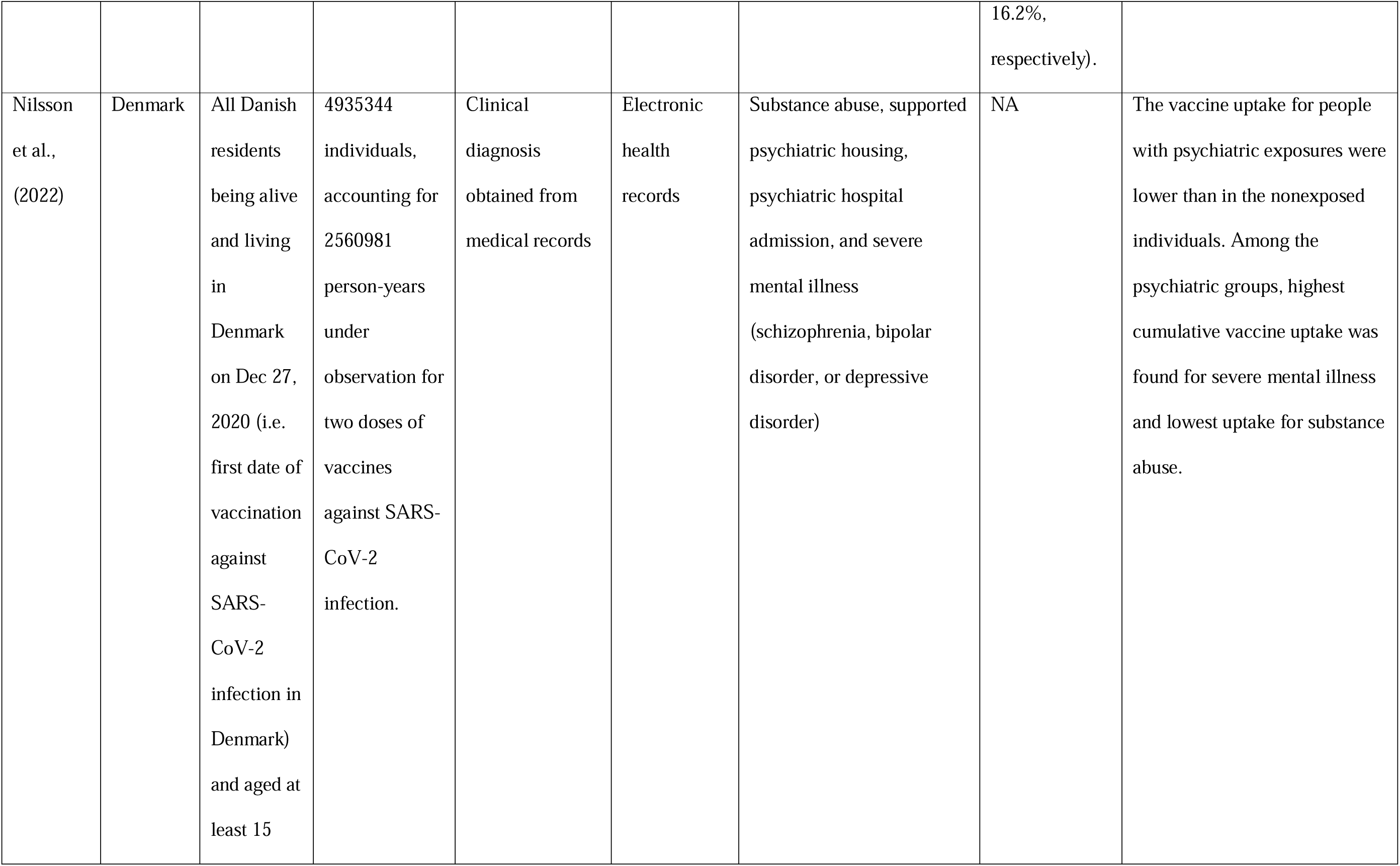
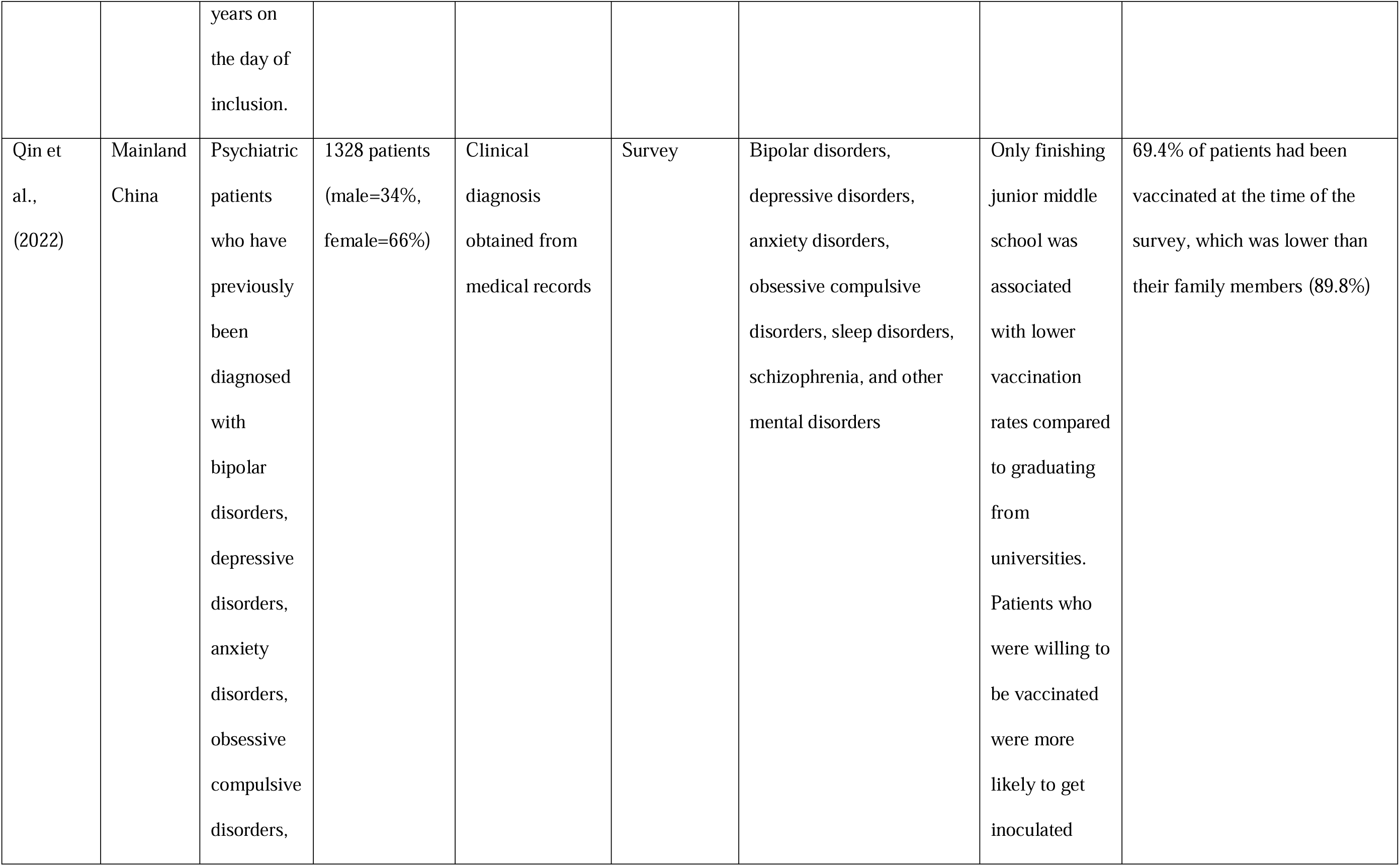
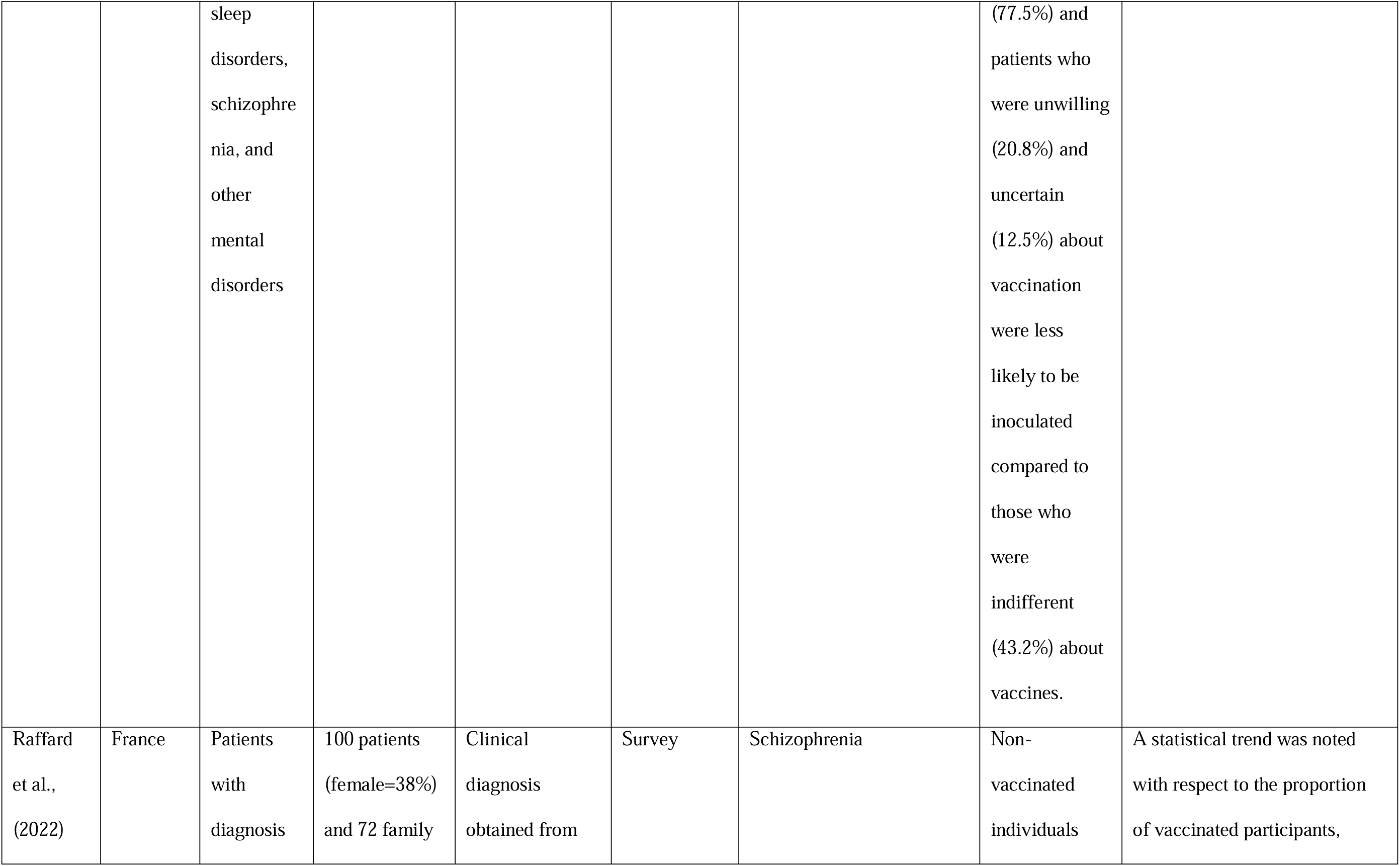
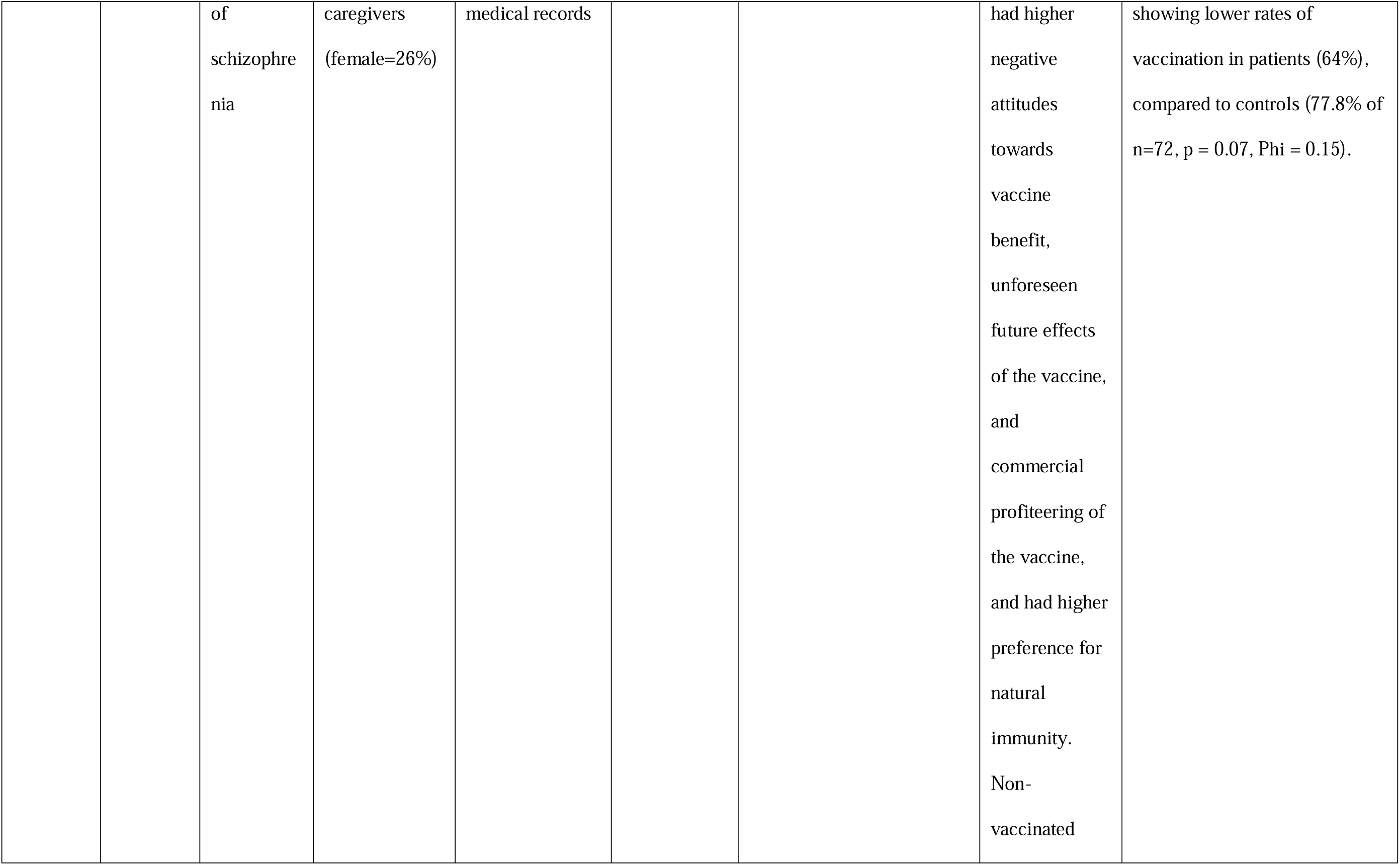
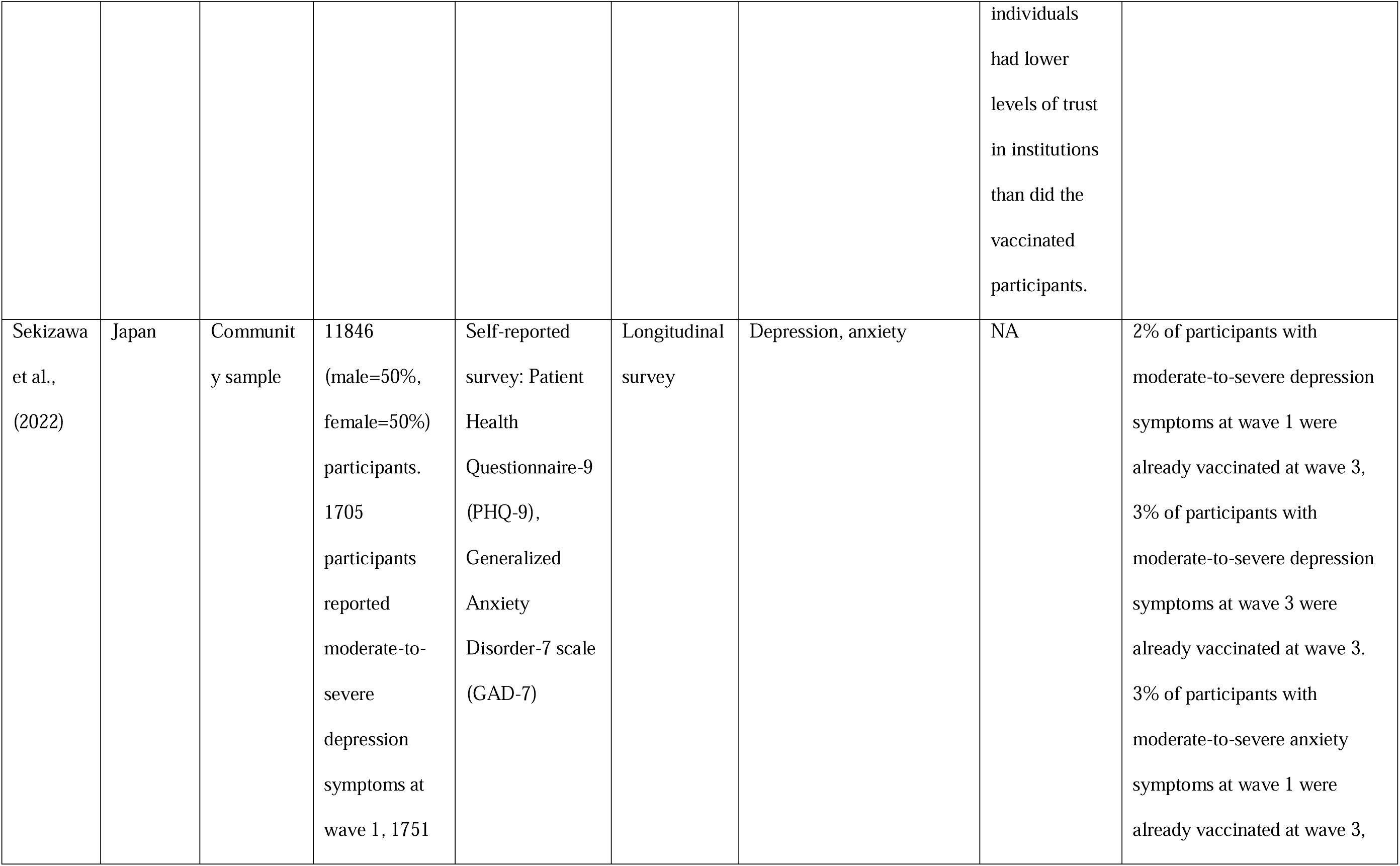
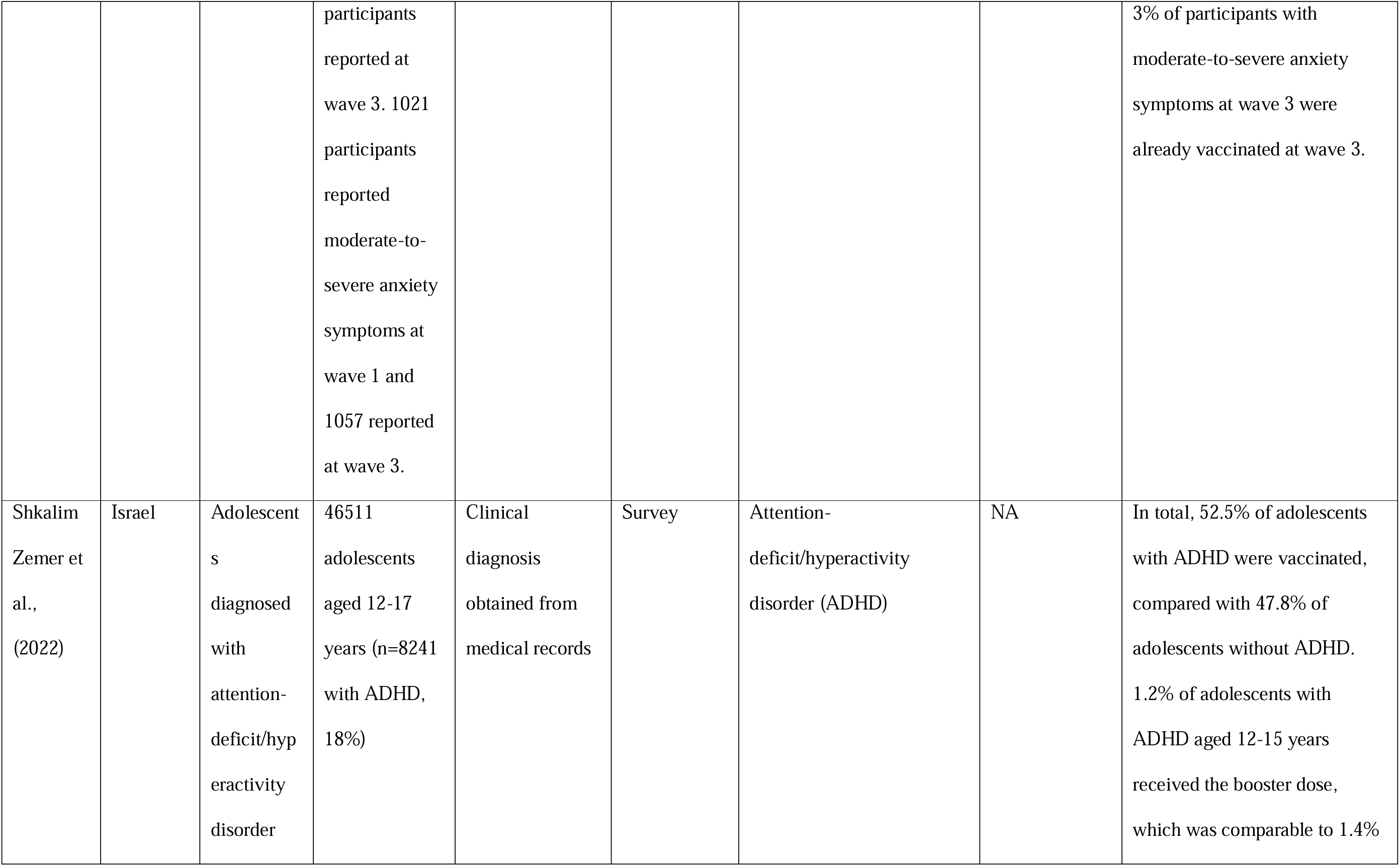
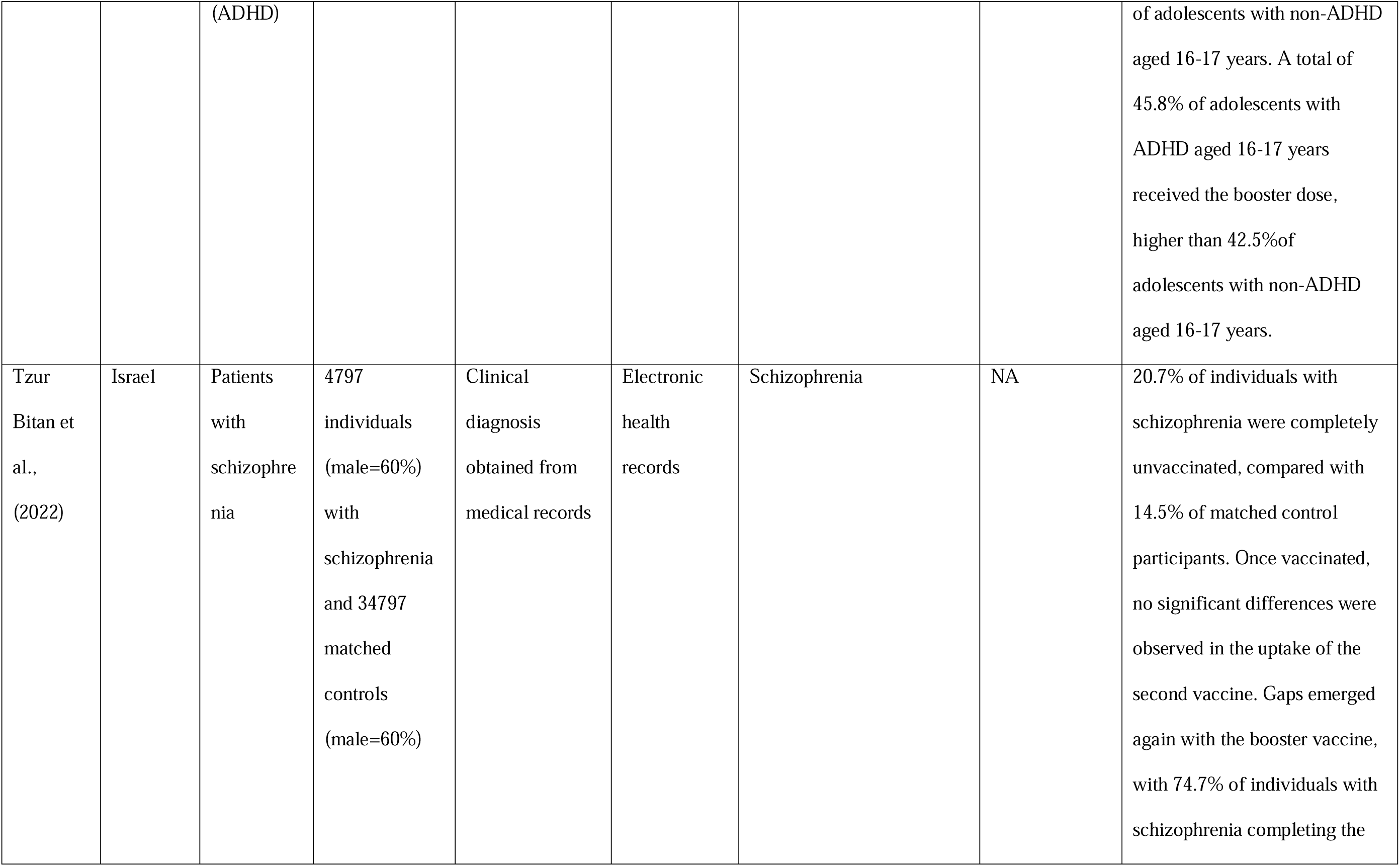
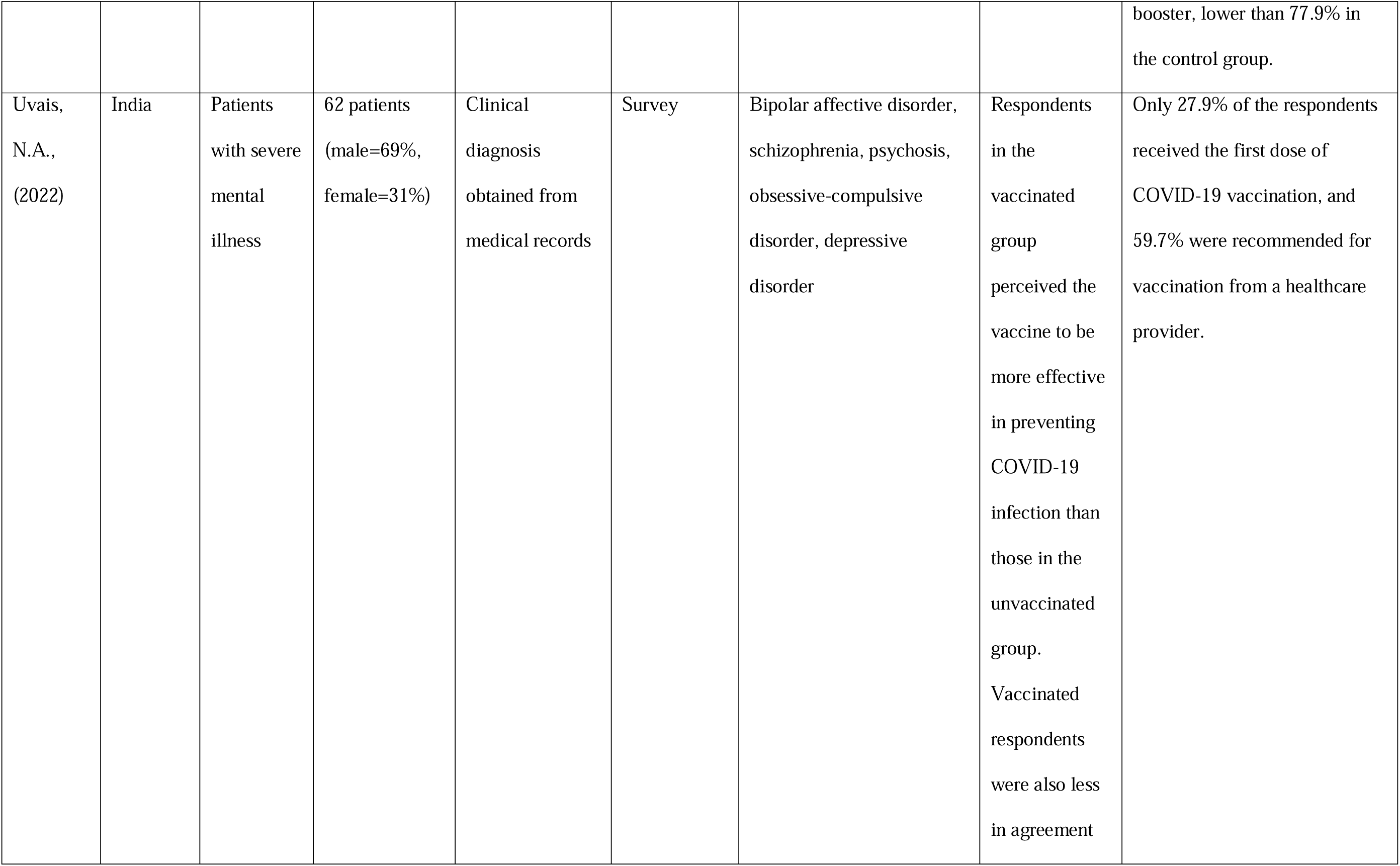
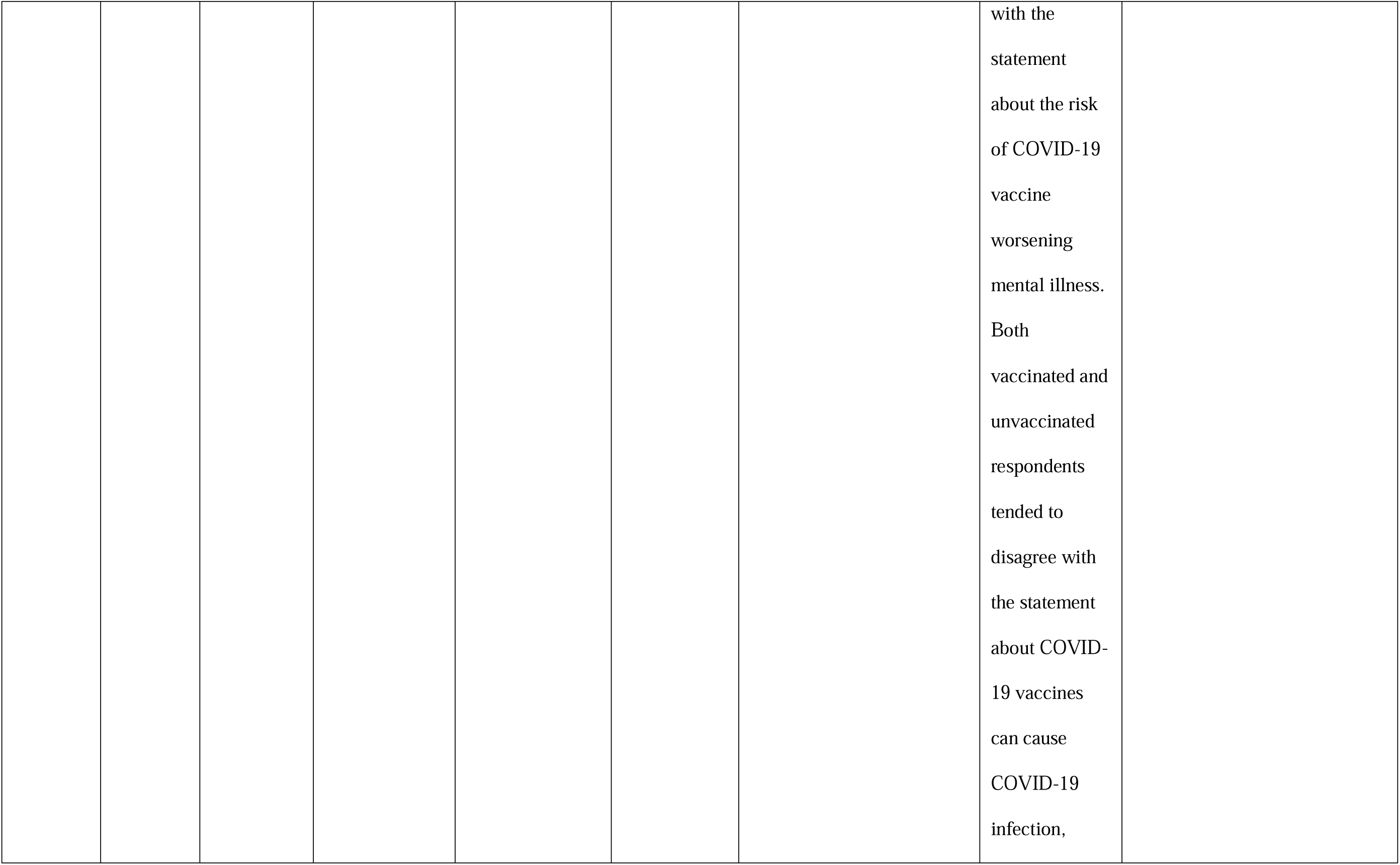
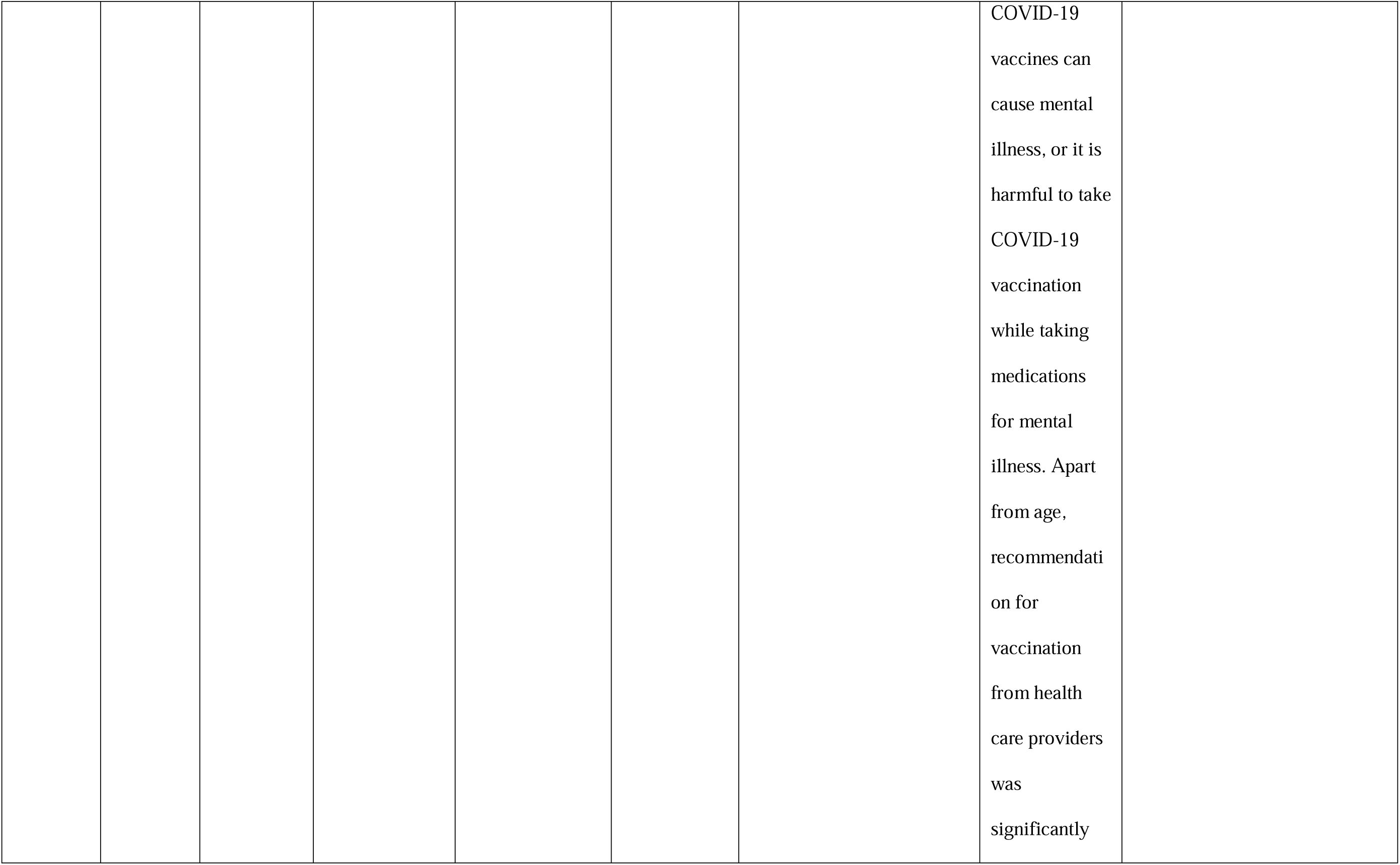
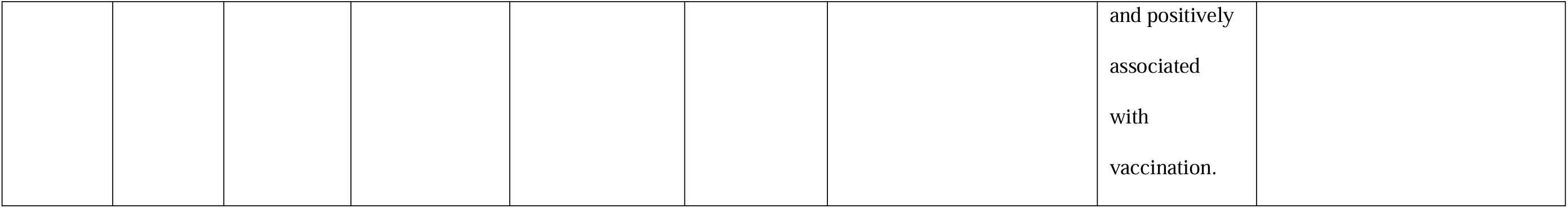
Summary of studies investigating the association between mental health conditions and COVID-19 vaccine uptake.

In the 17 studies, the reported uptake of COVID-19 vaccines (including boosters) ranged from 2% to 93%, among patients with mental health conditions. One study was conducted early in the pandemic when the COVID-19 vaccines were not widely available (October 2020 – May 2021), thus their reported COVID-19 vaccine uptake rates were low (2-3%, Sekizawa et al., 2022). After excluding this study, other studies reported an uptake of 11-93% among patient with mental health conditions.

#### Differences between people with and without mental health conditions

Thirteen of the 17 studies compared COVID-19 vaccine uptake among people with mental health conditions with uptake among people without these conditions. Eight of these 13 studies observed that uptake was lower in people with mental health conditions (Curtis, Inglesby, MacKenna, et al., 2022; Curtis, Inglesby, Morton, et al., 2022; Murphy et al., 2022; Nguyen et al., 2022; Nilsson et al., 2022; Qin et al., 2022; Raffard et al., 2022; Tzur Bitan et al., 2022). The reported vaccine uptake was between 3-21% lower among people with mental health conditions. Five of the 13 studies observed that the vaccine uptake was up to 1.19 times higher among patients with mental health conditions, compared with people without (Balut et al., 2021; Hassan et al., 2022; Mazereel et al., 2021; Moeller et al., 2021; Shkalim Zemer et al., 2022). However, among these five studies, two further reported significantly higher rates of declining the COVID-19 primary or booster vaccines among people with mental health conditions, compared with people without (Hassan et al., 2022; Shkalim Zemer et al., 2022). Two of the 17 studies reported the uptake of booster vaccines. Shkalim Zemer et al., (2022) reported higher uptake of the booster vaccine among adolescents with ADHD aged 16-17 (46%) compared with non-ADHD adolescents of the same age (43%), and similar uptake of the booster vaccine among adolescents with ADHD aged 12-15 and non-ADHD adolescents of the same age (1.2% vs 1.4%). Tzur Bitan et al., (2022) reported significantly lower booster vaccine uptake among patients with schizophrenia (75%) compared with people without (78%).

#### Differences between different mental health conditions

Two of the 17 studies compared vaccine uptake across different psychiatric diagnoses but reported different observations. One reported that 51% of patients with schizophrenia were vaccinated, which was lower (significance levels unreported) than those with other psychiatric disorders: bipolar disorder, 74%; depression disorder, 74%; anxiety disorder, 66%; obsessive-compulsory disorder, 85%; sleep disorder, 80%; other psychiatric disorders, 65% (Qin et al., 2022). Another study, however, did not find significant effects of psychiatric diagnosis on vaccine uptake (Mazereel et al., 2021).

#### Barriers to COVID-19 vaccine uptake among people with mental health conditions

We also extracted results on barriers to the COVID-19 vaccine uptake among people with mental health conditions from these studies. Five of the 17 studies reported results on this (Huang et al., 2021; Nguyen et al., 2022; Qin et al., 2022; Raffard et al., 2022; Uvais, 2022). Four of the five studies reported that, among people with mental health conditions, unvaccinated individuals were more likely to report greater concerns about the safety, effectiveness, and side effects of the COVID-19 vaccine than vaccinated people (Huang et al., 2021; Nguyen et al., 2022; Raffard et al., 2022; Uvais, 2022). Lack of trust in the vaccine, development of the vaccine, and the government was reported among unvaccinated people with mental health conditions in two studies (Nguyen et al., 2022; Raffard et al., 2022). One study reported that the vaccination rate was higher among patients who had the intention to be vaccinated (78%), compared with those who were unwilling to be vaccinated (21%) or those who were uncertain about the vaccination (13%, Qin et al., 2022). One study reported that recommendation for vaccination from health care providers was positively associated with vaccine uptake in people with mental health conditions (Uvais, 2022).

Overall, the observed uptake of COVID-19 vaccines among people with mental health conditions varied between studies. The results were mixed, with 8/13 studies reporting lower vaccine uptake in people with mental health conditions and 5/13 studies reporting no differences. Although with limited evidence, patients with schizophrenia might have lower COVID-19 vaccine uptake, compared with people without mental health conditions and people with other mental health conditions. Among people with mental health conditions, the most common barriers to COVID-19 vaccine uptake include concerns about the safety, effectiveness, and side effects of the COVID-19 vaccine.

#### Mental health conditions and COVID-19 vaccine breakthrough

Only two out of 33 studies reported outcomes related to breakthrough infection following vaccination in people with mental health conditions (Table 3, Nishimi, Neylan, et al., 2022; Wang et al., 2022). Both studies defined a COVID-19 vaccine breakthrough infection as *“a positive SARS-CoV-2 test record occurring in the medical records* ≥*14 days after their final SARS-CoV-2 vaccine dose”*, in line with the CDC definition (Birhane et al., 2021). Wang et al., (2022) included patients with current substance use disorders, while Nishimi, Neylan, et al., (2022) included patients who had psychiatric disorder diagnoses within the past five years. The observation period was from February 2020 to November 2021 in Nishimi, Neylan, et al., (2022), and from December 2020 to August 2021 in Wang et al. (2022).

**Table 3.**
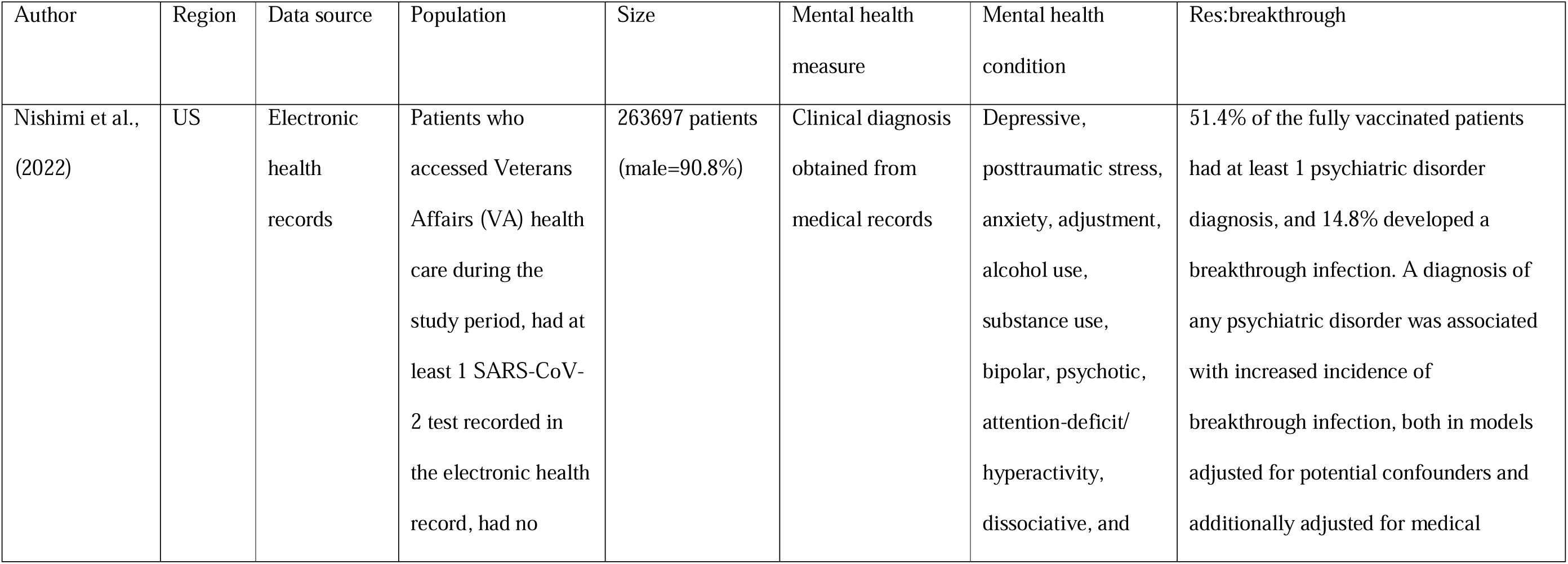
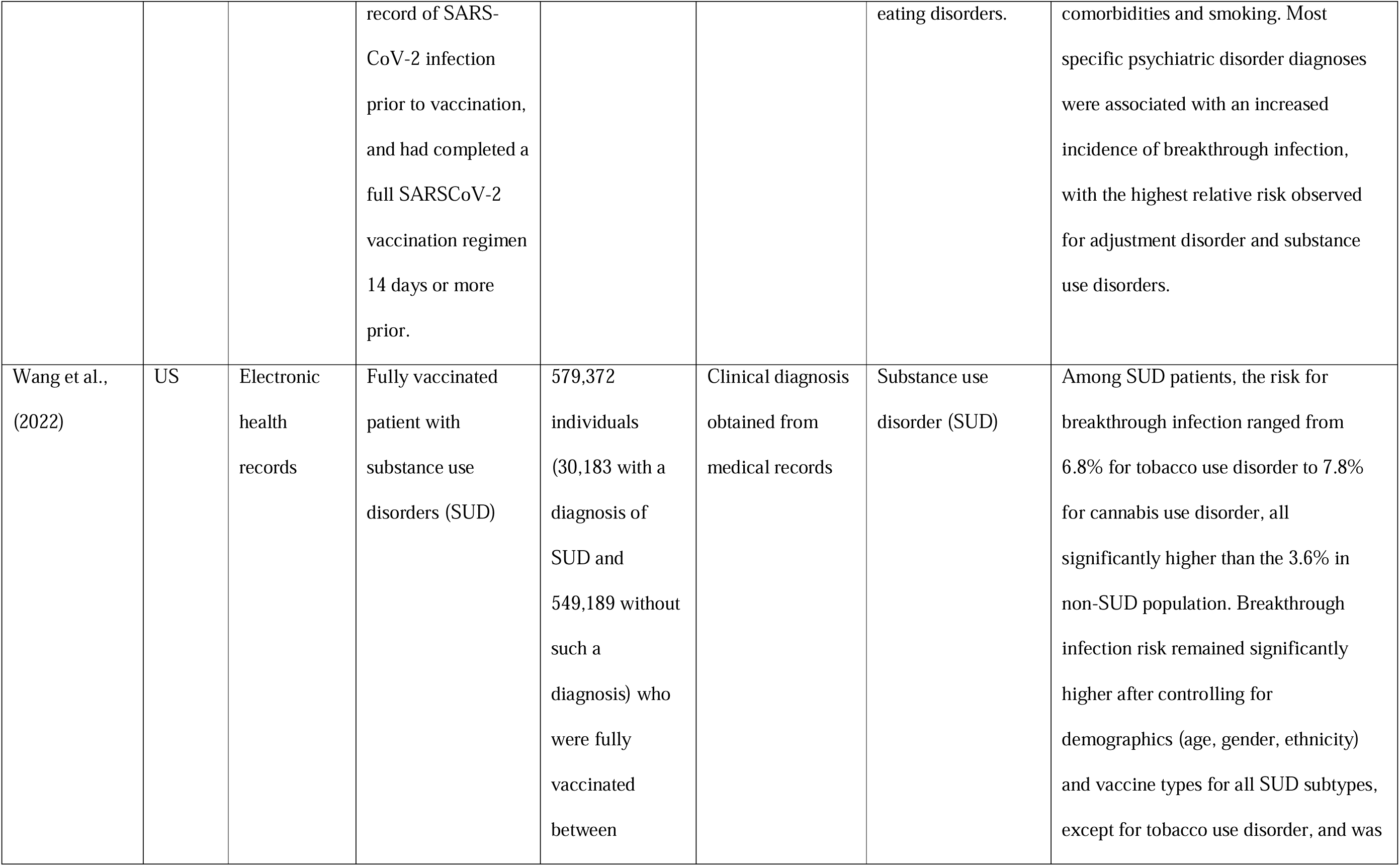
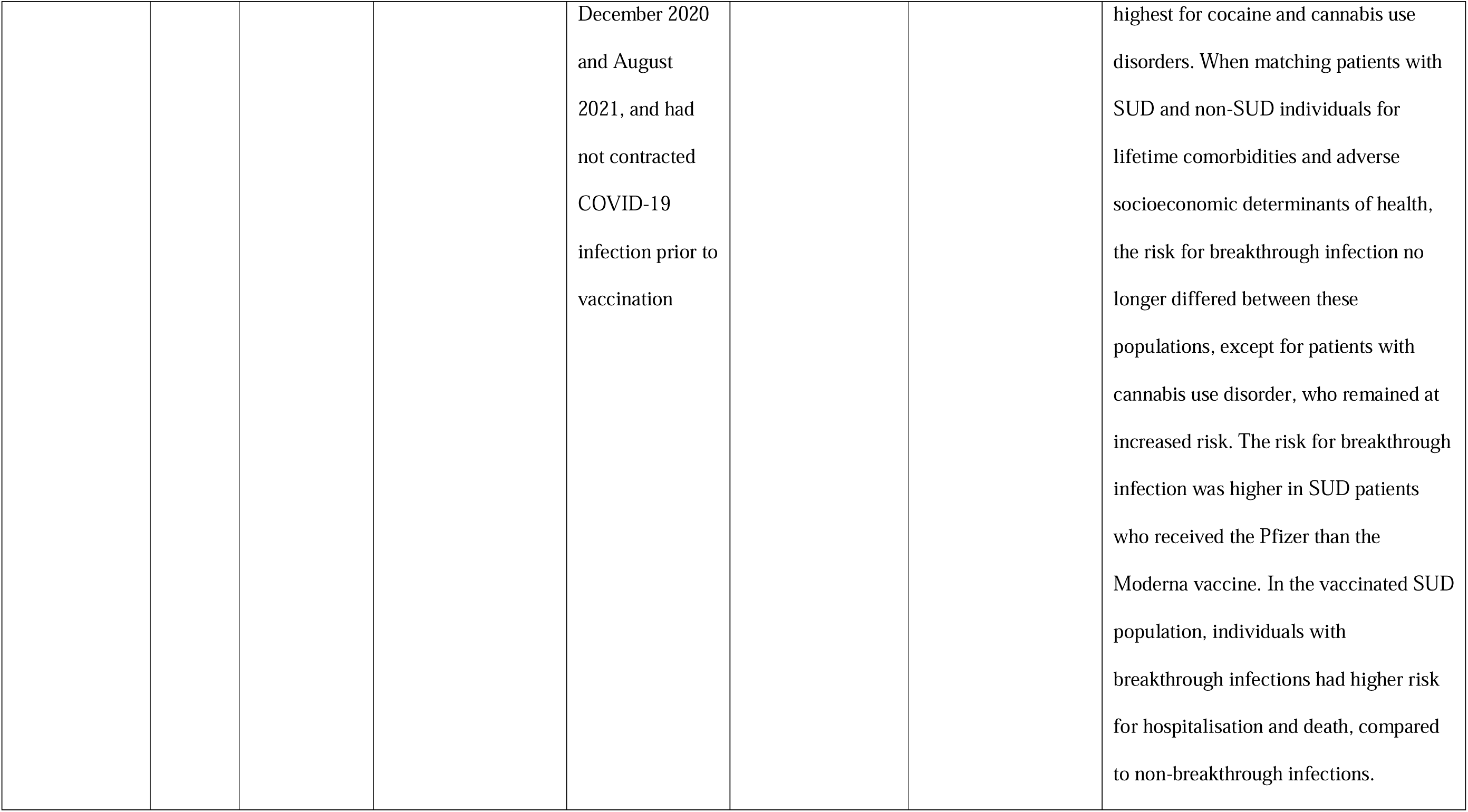
Summary of studies investigating the association between mental health conditions and COVID-19 vaccine breakthrough.

Both studies reported that people with mental health conditions were at increased risk of breakthrough infections for COVID-19. Specifically, Nishimi, Neylan, et al., (2022) reported that patients with any psychiatric disorder were 7% more likely to experience a breakthrough infection (95% CI, 1.05-1.09; *P*L<L.001), compared with patients without psychiatric disorders, after adjusting for potential confounders (e.g., age, ethnicity, vaccine type, time since vaccine, etc.). These patients were still at 3% increased risk of experiencing breakthrough infection (95% CI, 1.01-1.05; *P*L<L.001), compared with patients without psychiatric disorders, after further adjusting for medical comorbidities (e.g., diabetes, cardiovascular diseases etc,) and smoking status. Nishimi, Neylan, et al., (2022) also reported that when considering specific psychiatric disorder diagnoses (i.e., major depressive disorder, PTSD, anxiety disorder, adjustment disorder, alcohol use disorder, substance use disorder, bipolar disorder, and other psychiatric disorder) the highest relative risk was observed for adjustment disorder (relative risk= 1.13; 95% CI: 1.10-1.16, *P*L<L.001) and substance use disorders (relative risk=1.16; 95% CI: 1.12-1.21, *P*L<L.001), after adjusting for confounders, medical comorbidities, and smoking status and there were no significant increases for psychotic disorder (relative risk=1.05; 95% CI, 0.99-1.11; *P*L=L.09).

The other study, Wang et al., (2022), reported that among patients with substance use disorders, the risks for breakthrough infections were 7% for tobacco use, 7% for opioid use, 7% for alcohol use, 8% for cocaine use, and 8% for cannabis use, all significantly higher than that in matched cohorts without substance use disorder (2-4%, *P*<0.001). The modest but higher risks of breakthrough infection among patients with substance use disorder remained significant after controlling for age, gender, ethnicity and vaccine types, for all but tobacco use disorder. However, after controlling for socioeconomic determinants of health (e.g., problems related to education, employment, occupational risk factors, and housing and economic circumstances) and physical comorbidities, the risks for breakthrough infection no longer differed between patient with substance use disorder and matched cohorts for all but cannabis use disorder. Patients with cannabis use disorder remained at increased risk (hazard ratio=1.55, 95% CI: 1.22-1.99) compared with matched cohorts without cannabis use disorder. When looking at severe outcomes of vaccine breakthrough infections (i.e., COVID-19 hospitalisation and death), the risk of hospitalization was 22.5% for patients with substance use disorders who experienced breakthrough infections and 1.6% for patients without breakthrough infections. The risk of death was 1.7% for patients with substance use disorders who experienced breakthrough infections and 0.5% respectively. When looking at the type of vaccines patients with substance use received, the risk for breakthrough infection was higher in patients who received the Pfizer than the Moderna vaccine (hazard ratio=1.49, 95% CI: 1.31-1.69).

Findings from these two studies suggest that patients with mental health conditions, especially those with substance use disorder, might be at a slightly increased risk of experiencing breakthrough infection after being vaccinated against COVID-19.

## Discussion

This review highlights the evidence and gaps in research concerning mental health conditions and three COVID-19 vaccine outcomes: vaccine intentions, vaccine uptake, and breakthrough infection. Overall, the evidence was mixed regarding the level of intention to accept the COVID-19 vaccine among individuals with mental health conditions, compared with those without. This finding largely agrees with previous reviews, where a scarcity of evidence of vaccine hesitancy among people with mental health conditions was reported (Kafadar et al., 2022; Payberah et al., 2022). This is possibly due to the levels of vaccine hesitancy or willingness varying across different mental health conditions, which was not considered in previous reviews. In this scoping review, we further differentiated the levels of vaccine hesitancy between different mental health conditions. We found evidence reporting that patients with substance use disorders had higher odds for vaccine hesitancy, while patients with bipolar disorder had lower odds for vaccine hesitancy (Eyllon et al., 2022). Another study included in this review also reported that community-dwelling patients had a higher proportion of COVID-19 vaccination hesitancy compared to hospitalised patients (Bai et al., 2021). Therefore, combining people with different mental health conditions, or combining hospitalised patients and community-dwelling patients together in such investigations might lead to different results. Future research should divide patients into subgroups of different mental health conditions or diagnoses to understand the association between different conditions or diagnoses and vaccine willingness and hesitancy.

### Intention versus behaviour

One unique contribution of this review is that we also summarised evidence on the actual uptake of COVID-19 vaccines among people with mental health conditions, and did not rely on data on vaccine intentions. One of the reviewed studies reported results on the intention-behaviour association: the vaccination rate was higher among patients who were willing to be vaccinated compared with those who were unwilling or uncertain about the vaccination. This result implies that vaccine hesitancy (intention) is a valuable target for interventions to address low vaccine uptake. However, although tackling vaccine hesitancy among people with mental health conditions is important, it might not be sufficient to encourage vaccine uptake in this population. According to the Theory of Planned Behaviour (TPB), attitude towards the behaviour, subjective norm, and perceived behavioural control are three determinants of a behavioural intention, which then lead to a behaviour. The behavioural intention is an “immediate antecedent of behaviour” (Ajzen & Manstead, 2007). Therefore, although intention is a key contributor to behaviour, it might not necessarily translate into a certain behaviour directly. A meta-analysis on the efficacy of TPB examined to what extent behavioural intentions predicted actual behaviours (Armitage & Conner, 2001). They reported that, intention accounted for only 25% variance in behaviours, and the correlation between intention and behaviour was 0.47. This intention-behaviour discordance has been reported in previous literature and in other contexts. For example, a meta-analysis focusing on physical activity found that 36% of people who intended to engage in physical activity failed to do so (Rhodes & de Bruijn, 2013). In a more recent study where TPB was used as the theoretical model to explore the associations between intention and the behaviour of receiving Human papillomavirus (HPV) vaccines using TPB, intention accounted only for 9.6% of the variance in behaviour, while moral norm (i.e., the perceived moral correctness of a behaviour) and intention together accounted for 14.1% of the variances in behaviour (Juraskova et al., 2012).

In the context of vaccine hesitancy, it was also pointed out in literature that there might be circumstances where vaccine hesitancy is not the sole or main contributor to low vaccine uptake (MacDonald, 2015). In this review, the evidence on COVID-19 vaccine intentions and uptake among people with mental health conditions also seems to suggest a discordance between the two. Although the reported uptake of the COVID-19 vaccine varied across studies, depending on the timings, settings, and populations under investigation, most of the studies suggested that people with mental health conditions were more likely to decline the vaccine when offered. Especially when looking at studies (k=8 of 17) using electronic health records with large sample sizes (n>4000), the reported rate of COVID-19 vaccine uptake among people with mental health conditions ranged from 47% to 90% (Balut et al., 2021; Curtis, Inglesby, MacKenna, et al., 2022; Curtis, Inglesby, Morton, et al., 2022; Hassan et al., 2022; Murphy et al., 2022; Nilsson et al., 2022; Shkalim Zemer et al., 2022; Tzur Bitan et al., 2022). Five out of these eight studies reported that people with mental health conditions had significantly lower uptake of the COVID-19 vaccine, compared with people without mental health conditions (Curtis, Inglesby, MacKenna, et al., 2022; Curtis, Inglesby, Morton, et al., 2022; Murphy et al., 2022; Nilsson et al., 2022; Shkalim Zemer et al., 2022; Tzur Bitan et al., 2022), while three reported the opposite (Balut et al., 2021; Hassan et al., 2022; Shkalim Zemer et al., 2022). The nature of this intention-behaviour gap, and other barriers (e.g., other systematic-level barriers) that contribute to low vaccine uptake, need further understanding. Interventions targeting at other aspects of vaccine delivery may also be considered, for example, making vaccines available at psychiatric clinics, providing psychiatrists with sufficient information about the vaccines, providing more support for patients with mental health conditions to receive vaccines at GPs, having engagement campaigns and activities tailoring to people with mental health conditions and their carers, etc (Mazereel et al., 2021; Payberah et al., 2022).

### Understanding barriers to vaccine hesitancy and uptake in people with mental health conditions

According to the WHO’s “3Cs” model of vaccine hesitancy, confidence, complacency, and convenience are the three main determinants of vaccine hesitancy (Sage Working Group on Vaccine Hesitanc, 2014). The “confidence” element mainly includes trusts in the vaccines (e.g., safety, effectiveness, the delivery system, the government, etc.). The “complacency” element mainly includes the perceived risks of vaccine-preventable diseases. Both the “confidence” and “complacency” elements reflect more on the individual-level barriers. The “convenience”, on the other hand, mainly reflects systematic-level barriers around the availability, affordability, and accessibility of the vaccine and vaccine services. Both individual-level barriers and system-level barriers may contribute to the lower vaccine uptake in people with mental health conditions (Warren et al., 2021). We also summarised findings regarding these barriers. Our findings suggest that people with mental health conditions are more likely to experience barriers related to the “confidence” (i.e., concerns about the safety, effectiveness, and side effects of the COVID-19 vaccine) and “convenience” (e.g., whether people with mental health conditions received recommendations from health care providers) of the COVID-19 vaccines. These findings suggest studies and interventions should aim to tackle both the individual-level and system-level barriers, especially targeting vaccine “confidence” and “convenience”, to encourage the uptake of vaccines in people with mental health conditions. For example, outreach by mental health care providers may be a strategy to improve COVID-19 vaccine uptake in this population. A pilot trial found that when psychiatric providers were actively engaged in addressing concerns about the COVID-19 vaccine with patients with severe mental illness during outpatient visits, the vaccination rate among outpatients was higher (84%) than estimated (62.1% - 77.3%; Lim et al., 2022). Other strategies such as identifying leaders in the communities to promote vaccination outreach (Sullivan et al., 2022), transparent communication concerning the safety and effectiveness of the COVID-19 vaccines (Eyllon et al., 2022; Jefsen et al., 2021; Sullivan et al., 2022) may also be helpful. Another observation from this review was that, two of the five studies reporting higher or comparable vaccine uptake in people with mental health conditions were conducted among adolescents or children (Moeller et al., 2021; Shkalim Zemer et al., 2022), while all other studies were conducted in adult populations. It is possible that their caregivers’ opinions determined vaccine uptake in this group (Shkalim Zemer et al., 2022). Caregiver-related barriers and facilitators to vaccine uptake should also be considered in future studies investigating vaccine uptake in people, especially children or adolescents, with mental health conditions.

### Vaccine breakthrough in people with mental health conditions

Another contribution of this review is the summary on the association between mental health conditions and COVID-19 vaccine breakthrough. Only two published studies were identified and they both suggested that vaccine breakthrough may be a particular risk for those with substance use disorder (Nishimi, Neylan, et al., 2022; L. Wang et al., 2022). Wang et al. (2022) further reported that these patients were also at risk of more adverse outcomes, including hospitalisation and death, following a breakthrough infection, compared with non-breakthrough infections. These results suggest that psychological stressors may influence the susceptibility to COVID-19. However, both studies were conducted in the US. There is no other evidence that can support generalisability of these findings in other countries and populations. In general, findings from both studies echo evidence from prior to the COVID-19 pandemic that both psychological and social factors affect vulnerability to viral infections and also vaccine effectiveness. For example, the series of viral challenge studies by Sheldon Cohen and colleagues, where they found that individuals with higher levels of psychological stress or experiencing chronic stressors were at significantly increased risk of acute infectious respiratory illness when exposed to respiratory viruses (e.g., rhinovirus type 2, 9, or 14, respiratory syncytial virus, coronavirus type 229E, rhinovirus RV39, Hanks; Cohen et al., 1991, 1998). Other observational and interventional studies have also shown a consistent pattern in which poorer mental health (e.g., exposure to more stressful life events, bereavement, chronic stress) is associated with sub-optimal vaccine responses (Gallagher et al., 2008; Phillips et al., 2006; Vedhara et al., 1999). The evidence together shows that mental health is associated with an increased risk of illness from viral infection, and increased risk of impaired protection following vaccination over time. This again emphasises the need to encourage people with mental health conditions to receive vaccinations to protect them from increased risks of infections and subsequent illnesses. However, there is need to investigate the risks of vaccine breakthrough in people with mental health conditions in other countries or regions, to allow comparison of the impact of different healthcare systems.

## Limitations

This scoping review did not assess the quality of the studies or quantitatively assess the relationships between mental health and COVID-19 vaccine outcomes using meta-analysis. For studies concerning COVID-19 vaccine willingness or hesitancy, settings (e.g., in hospital or in community) and populations varied between studies which may limit the generalisability of the findings. In addition, 90% of these studies measured vaccine willingness or hesitancy using different self-reported item(s) where self-report biases may be present (Johnson & Fendrich, 2005). For studies concerning COVID-19 vaccine uptake and breakthrough, studies did not distinguish between people who were experiencing current mental health conditions and who had previous mental health diagnosis. It is unclear whether current or previous mental health conditions would have any impact on vaccine uptake or breakthrough. Only two studies reported results concerning COVID-19 vaccine breakthrough among people with mental health conditions, suggesting that evidence on this topic is currently limited.

## Future directions

Urgent research effort is needed to investigate the risks of COVID-19 vaccine breakthrough and severe COVID-19 outcomes among people with mental health conditions, and associated risk factors. Future studies should also be encouraged to distinguish between the different mental health conditions and also the impact of current or previous mental health conditions on COVID-19 vaccine outcomes. The nature of barriers to vaccine uptake, and the intention-behaviour gap in vaccinations also need further investigation.

## Conclusions

This is the first review, to our knowledge, focusing on the three COVID-19 vaccine outcomes (i.e., intention, uptake, and vaccine breakthrough) among people with mental health conditions. We found that the evidence regarding the level of vaccine intentions in this population was inconclusive. However, the majority of evidence seemed to suggest that the level of vaccine uptake among people with mental health conditions was lower than people without. There might be an intention-behaviour gap among people with mental health conditions, in the context of COVID-19 vaccination. Future studies may investigate both the nature of this intention-behaviour gap, and strategies to encourage vaccine uptake in this population. Limited evidence seemed to also suggest that people with mental health conditions may be at increased risk of COVID-19 vaccine breakthrough. Future work may also investigate the scale of this issue, and possible consequences.

## Data Availability

All data produced in the present work are contained in the manuscript.

